# Longitudinal analysis shows durable and broad immune memory after SARS-CoV-2 infection with persisting antibody responses and memory B and T cells

**DOI:** 10.1101/2021.04.19.21255739

**Authors:** Kristen W. Cohen, Susanne L. Linderman, Zoe Moodie, Julie Czartoski, Lilin Lai, Grace Mantus, Carson Norwood, Lindsay E. Nyhoff, Venkata Viswanadh Edara, Katharine Floyd, Stephen C. De Rosa, Hasan Ahmed, Rachael Whaley, Shivan N. Patel, Brittany Prigmore, Maria P. Lemos, Carl W. Davis, Sarah Furth, James O’Keefe, Mohini P. Gharpure, Sivaram Gunisetty, Kathy A. Stephens, Rustom Antia, Veronika I. Zarnitsyna, David S. Stephens, Srilatha Edupuganti, Nadine Rouphael, Evan J. Anderson, Aneesh K. Mehta, Jens Wrammert, Mehul S. Suthar, Rafi Ahmed, M. Juliana McElrath

**Affiliations:** Vaccine and Infectious Disease Division, Fred Hutchinson Cancer Research Center, Seattle, WA, 98109, USA; Emory Vaccine Center and Department of Microbiology and Immunology, Emory University, Atlanta, GA 30322, USA; Department of Medicine, Division of Infectious Diseases, Hope Clinic of Emory Vaccine Center, Emory University School of Medicine, Atlanta, GA 30329, USA; Center for Childhood Infections and Vaccines of Children’s Healthcare of Atlanta, Emory University Department of Pediatrics Department of Medicine, Atlanta, GA 30322, USA; Emory University School of Medicine, Department of Medicine, Atlanta, GA 30322, USA; Departments of Laboratory Medicine and Medicine, University of Washington, Seattle, WA 98195, USA; Department of Biology, Emory University, Atlanta, GA 30322, USA; Department of Microbiology and Immunology, Emory University, Atlanta, GA 30322, USA; Yerkes National Primate Research Center, Atlanta, GA 30329, USA

**Keywords:** SARS-CoV-2, COVID-19, Immune memory, Antibody, Kinetics, Neutralization, CD4+ T cells, CD8+ T cells, B cells, Spike, RBD, Endemic coronaviruses

## Abstract

Ending the COVID-19 pandemic will require long-lived immunity to SARS-CoV-2. Here, we evaluate 254 COVID-19 patients longitudinally up to eight months and find durable broad-based immune responses. SARS-CoV-2 spike binding and neutralizing antibodies exhibit a bi-phasic decay with an extended half-life of >200 days suggesting the generation of longer-lived plasma cells. SARS-CoV-2 infection also boosts antibody titers to SARS-CoV-1 and common betacoronaviruses. In addition, spike-specific IgG+ memory B cells persist, which bodes well for a rapid antibody response upon virus re-exposure or vaccination. Virus-specific CD4+ and CD8+ T cells are polyfunctional and maintained with an estimated half-life of 200 days. Interestingly, CD4+ T cell responses equally target several SARS-CoV-2 proteins, whereas the CD8+ T cell responses preferentially target the nucleoprotein, highlighting the potential importance of including the nucleoprotein in future vaccines. Taken together, these results suggest that broad and effective immunity may persist long-term in recovered COVID-19 patients.

## INTRODUCTION

The COVID-19 pandemic caused by the rapid spread of SARS-CoV-2, a novel betacoronavirus, continues to cause significant morbidity and mortality. The induction of effective early immune control of SARS-CoV-2 and durable immune memory is critical to prevent severe disease and to protect upon re-exposure. SARS-CoV-2 infection induces polyclonal humoral and cellular responses targeting multiple viral proteins described in cross-sectional and longitudinal studies.^1^ More comprehensive, quantitative analyses with extensive serial sampling in larger numbers of COVID-19 patients are limited, and could resolve some conflicting views about the durability of humoral immunity. Importantly, defining the frequency, immune function and specificity of the antibodies, memory B and T cell responses among COVID-19 patients, and identifying when they appear and how long they persist can provide understanding of the integral components for long-lived immunity to SARS-CoV-2 and potentially other human coronaviruses that emerge in the future.^2^

We initiated two prospective COVID-19 patient cohorts in Seattle and Atlanta during the first surge of the pandemic to investigate long-term immunity to SARS-CoV-2.

Among 254 COVID-19 patients enrolled and frequently sampled, we identify binding and neutralizing antibodies to SARS-CoV-2 as well as antigen-specific B and T cells elicited early after infection, define their specificities, quantify the extent of antibody boosting of cross-reactive responses to other coronaviruses, and further characterize the decay rate and durability of these immune parameters over 250 days. We employ highly standardized or validated assays that are also being used to evaluate immunity in recent and ongoing clinical vaccine trials.^3–5^ This indepth longitudinal study demonstrates that durable immune memory persists in most COVID-19 patients, including those with mild disease, and serves as a framework to define and predict long-lived immunity to SARS-CoV-2 after natural infection. This investigation will also serve as a benchmark for immune memory induced in humans by SARS-CoV-2 vaccines.

## RESULTS

### COVID-19 study population

COVID-19-confirmed patients were recruited into our longitudinal study of SARS-CoV-2 specific B and T cell memory after infection. A total of 254 patients enrolled at two sites, Atlanta and Seattle, starting in April 2020 and returned for follow up visits over a period of 250 days. We were able to collect blood samples at 2-3 time points from 165 patients and at 4-7 time points from another 80 patients, which allowed us to perform a longitudinal analysis of SARS-CoV-2-specific B and T cell responses on a large number of infected patients. The demographics and baseline characteristics of this cohort are described in Table S1. The study group was 55% female and 45% male and between 18-82 years old (median, 48.5 years). Based on World Health Organization (WHO) guidelines of disease severity, 71% of study participants exhibited mild disease, 24% had moderate disease, and 5% experienced severe disease.

### Antibody responses to SARS-CoV-2 spike protein show a bi-phasic decay with an

Binding antibodies to the SARS-CoV-2 full length spike protein, to the receptor binding domain (RBD) and to the N terminal domain (NTD) of the spike protein were assessed in COVID-19 patients (n=222) over a period of 8 months post symptom onset. We included healthy individuals age 18-42 years as negative controls whose longitudinal blood samples were collected before the emergence of the COVID-19 pandemic. These pre-pandemic samples (n=51) were from recipients of either the seasonal inactivated influenza vaccine (n=27, collected from 2014-2018) or the live yellow fever virus (YFV-17D) vaccine (n=24, collected from 2005-2007). The Mesoscale multiplex assay was used to measure IgG, IgA, and IgM antibody responses to SARS-CoV-2 proteins in the COVID-19 patients and in the pre-pandemic healthy controls.

The magnitude of serum IgG antibodies binding to the SARS-CoV-2 spike protein increased in 92% of COVID-19 convalescent participants (n=222) relative to pre-pandemic controls (Figure 1A). The IgG responses to SARS-CoV-2 spike, RBD, and NTD declined over time with half-lives of 126 (95% confidence interval [95%CI] [107, 154]), 116 (95%CI [97, 144]), and 130 (95%CI [110, 158]) days, respectively, as estimated by an exponential decay model (Figure 1A-C and S1A). We also estimated antibody waning using a power law model, which models a scenario in which the rate of antibody decay slows over time. The power law model produced a better fit for the decay of the SARS-CoV-2 spike, RBD, and NTD binding IgG antibodies (ΔAICs>10), Δ suggesting that spike-specific antibodies plateau over time. Because the decay rate changes over time, the half-life is predicted to change over time as well; therefore, we used the power law model to estimate the half-lives at 120 days after symptom onset.

**Figure 1.**
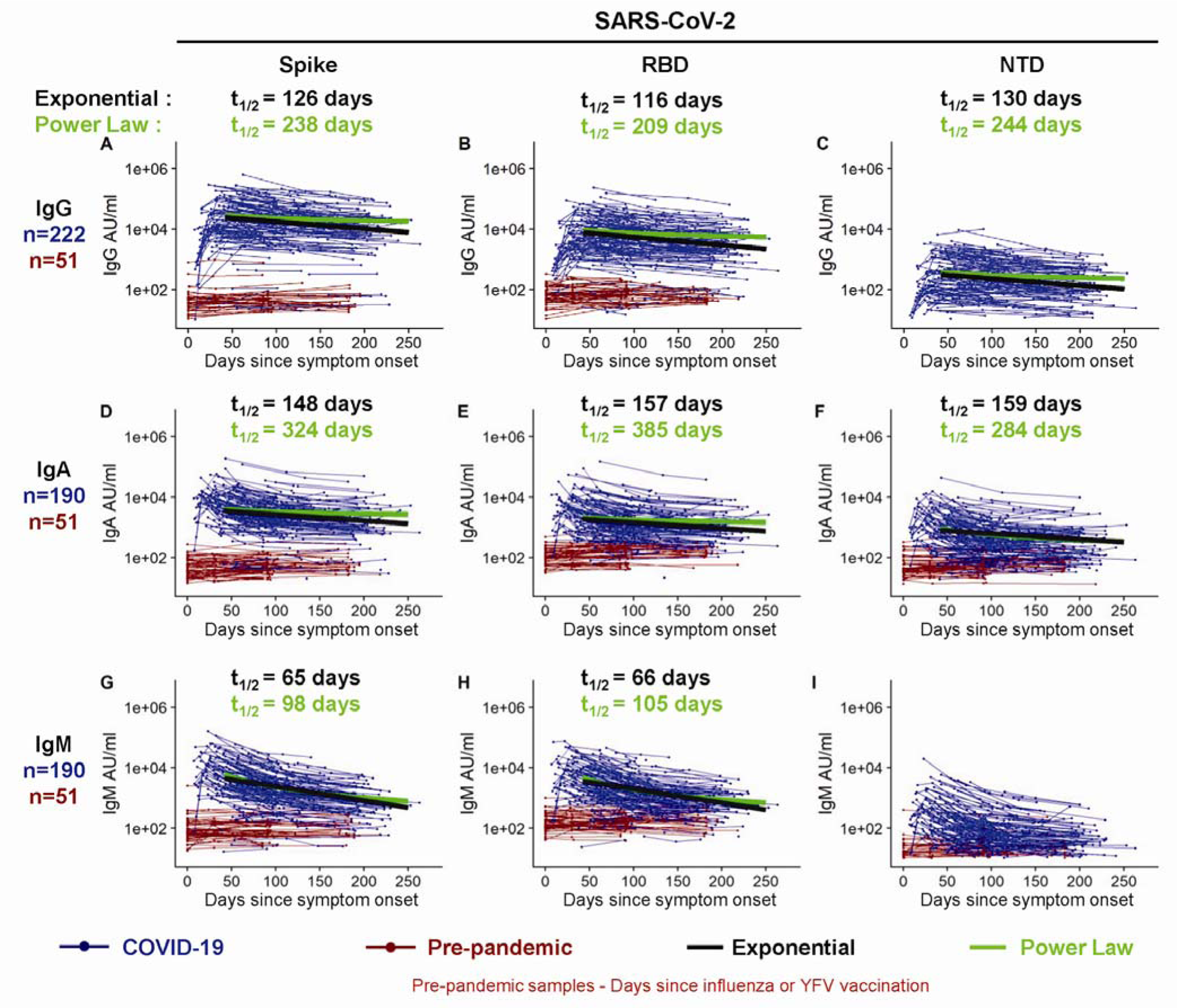
Longitudinal SARS-CoV-2 spike binding antibody responses. IgG (A-C), IgA (D-F), and IgM (G-I) antibodies reactive to SARS-CoV-2 spike (A, D, G), spike receptor binding domain (RBD; B, E, H), and the spike N terminal domain (NTD; C, F, I) were measured in triplicate by an electrochemiluminescent multiplex immunoassay and reported as arbitrary units per ml (AU/ml) as normalized by a standard curve. Longitudinal antibody titers of COVID-19 patients (in blue, n=222 COVID-19+ for IgG; n=190 COVID-19+ for IgA and for IgM) are plotted over days since symptom onset, whereas longitudinal pre-pandemic donor samples (in red, n=51 for IgG, IgA and IgM) were collected in the course of a non-SARS-CoV-2 vaccine study before 2019 and plotted over days since immunization. IgG decay curves and half-lives estimated by an exponential decay model are shown in black, and the decay curves and half-lives at day 120 post symptom onset estimated by a power law model are shown in green.

The power law estimated half-lives for the IgG antibody responses to spike (t_1/2_=238 days), RBD (t_1/2_=209 days), and NTD (t_1/2_=244 days) were longer than those estimated by the exponential decay model (Figure S1A, C), indicating that the concentration of these IgG antibodies may be starting to stabilize. IgA (Figure 1D-F) and IgM (Figure 1G-I) antibodies reactive to the SARS-CoV-2 spike also increased after SARS-CoV-2 infection but were detected at lower levels and declined faster than the SARS-CoV-2-reactive IgG antibodies. As expected, spike-binding IgM decayed more rapidly than spike-binding IgA and IgG. Taken together these results show that antibody responses, especially IgG antibody, were not only durable in the vast majority of patients in the 250 day period but also that the bi-phasic decay curve suggests the generation of longer lived plasma cells producing antibody to the SARS-CoV-2 spike protein.

We also examined the antibody response to the SARS-CoV-2 nucleocapsid protein in these infected patients. As expected, the COVID-19 patients showed higher levels of antibody to the nucleocapsid protein compared to the pre-pandemic healthy controls (Figure S2). However, the nucleocapsid-specific antibodies declined with a much shorter half-life of 63 days (95%CI [58, 70]) compared to the spike protein antibodies (Figure S1A, C). Also, the nucleocapsid reactive IgG decay rate was best fit by the exponential model and not the power law model in contrast to what we observed with the spike IgG antibody decay rate (Figure S1A). Thus, the nucleocapsid reactive IgG not only declined much faster but also showed less evidence of stabilizing antibody levels consistent with a response driven disproportionately by short-lived antibody secreting cells – at least at this stage of the immune response.

### Stable and long-lived antibody responses to common human alpha- and betacoronaviruses in pre-pandemic healthy controls

We were interested in determining if SARS-CoV-2 infection had any effect on the levels of antibody to the circulating human alpha- and betacoronaviruses. As a prelude to this question, we first examined antibody levels to the spike protein of the two circulating alphacoronaviruses (229E and NL63) and the two betacoronaviruses (HKU1 and OC43) in our pre-pandemic samples. As shown in Figure 2, all 51 pre-pandemic samples had clearly detectable levels of IgG and IgA antibodies to the spike proteins of the four human coronaviruses. This is the expected result since seropositivity to these coronaviruses is very high in the adult population, but what was quite interesting was the remarkable stability of these antibody responses over a 200-day period in the pre-pandemic serum samples (shown as red lines in Figure 2). These were essentially flat lines with no decline in the antibody levels and question the prevailing belief that antibody responses to the endemic coronaviruses are short-lived.^6–8^ While some occasional boosting of these childhood-acquired coronavirus infections cannot be ruled out, these data showing such stable antibody titers are best explained by the persistence of long-lived plasma cells in the bone marrow many years after infection.^9–13^

**Figure 2.**
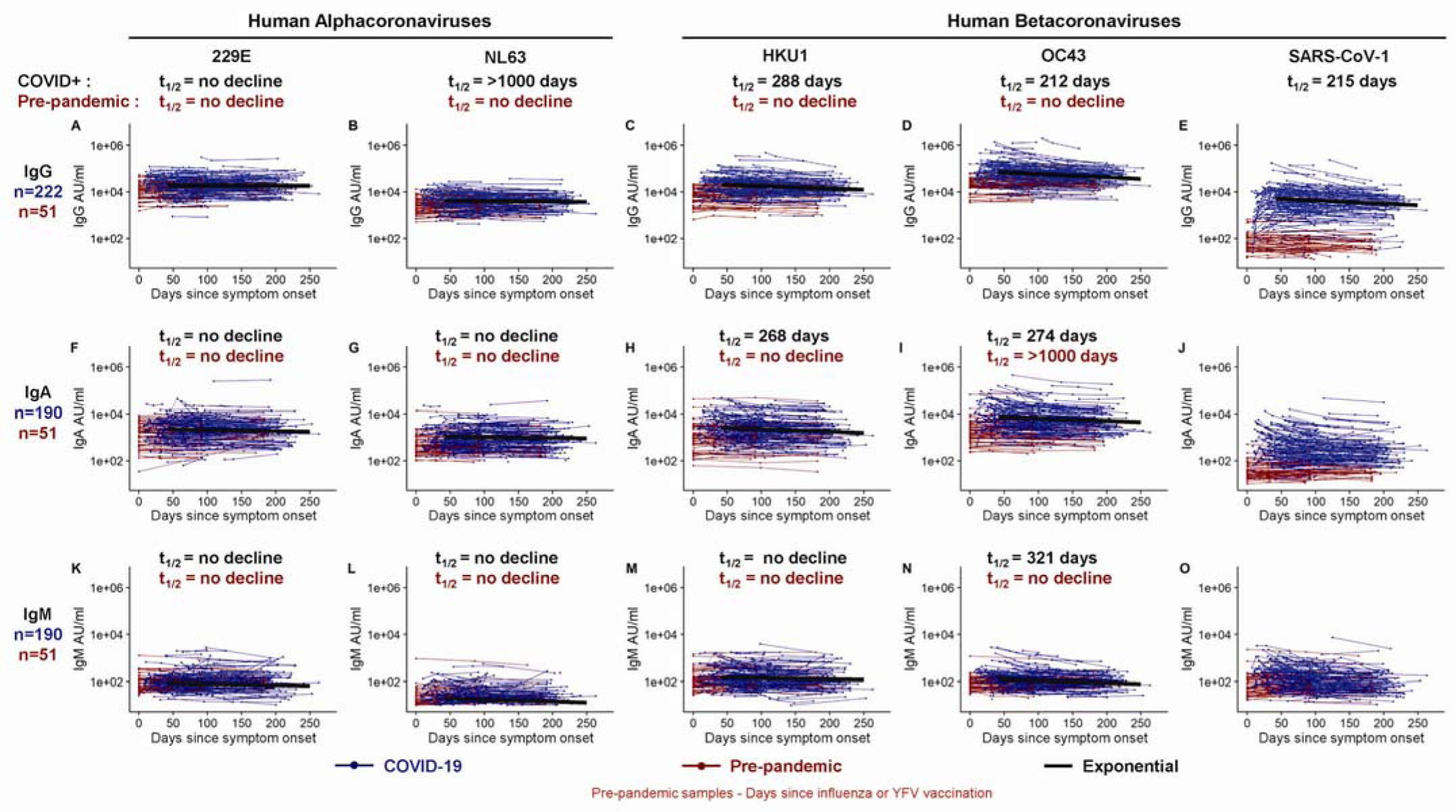
Longitudinal binding antibody responses to other coronavirus spike proteins. IgG (A-E), IgA (F-J), and IgM (K-O) antibody responses in sera collected from COVID-19+ patients (in blue, n=222 for IgG; n=190 for IgA and IgM) and pre-pandemic donors (in red, n=51 for IgG, IgA and IgM) that were measured to 229E spike (A, F, K), NL63 spike (B, G, L), HKU1 spike (C, H, M), OC43 spike (D, I, N), and the SARS-CoV-1 spike protein (E, J, O) in triplicate. Longitudinal antibody titers of COVID-19 patients are plotted over days since symptom onset, whereas longitudinal pre-pandemic donor samples were collected in the course of a non-SARS-CoV-2 vaccine study before 2019 and plotted over days since immunization. Antibody responses were measured by an electrochemiluminescent multiplex immunoassay and reported as arbitrary units per ml (AU/ml) as normalized by a standard curve. IgG decay curves and half-lives estimated by an exponential decay model are shown in black. There was no significant decline in IgG reactive to endemic alpha and betacoronaviruses in longitudinal samples collected in healthy donors before the pandemic (red, A-D).

### COVID-19 infection results in increased levels of antibodies to two common human betacoronaviruses (HKU1 and OC43) and to SARS-CoV-1

We next examined if SARS-CoV-2 infection had any impact on the levels of antibodies to the other human coronaviruses. We measured IgG, IgA, and IgM antibody binding to the spike proteins of other known human coronaviruses in the COVID-19 patients (n=222 for IgG and n=190 for IgA and IgM) and compared these data to the 51 pre-pandemic healthy donor samples. In the COVID-19 patients, IgG and IgA antibodies to the alphacoronaviruses, 229E and NL63, did not show any significant changes compared to the antibody levels in the pre-pandemic healthy controls (Figure 2A, B, F, G; Figure S1C, D). In contrast, the IgG and IgA antibodies to betacoronaviruses HKU1 and OC43 were substantially elevated in COVID-19 patients relative to pre-pandemic controls (Figure 2C, D, H, I; Figure S1C, D; p<0.0001). After this boost, HKU1 and OC43 IgG antibody levels declined with estimated half-lives of 288 (95%CI [235, 372]) and 212 (95%CI [176, 268]) days, respectively (exponential decay model). IgM levels to common betacoronaviruses HKU1 and OC43 were low in both pre-pandemic controls and COVID-19 patients (Figure 2M, N). While pre-existing exposure and antibodies against HKU1 and OC43 betacoronaviruses are common in adults, pre-existing SARS-CoV-1 exposure is rare and antibody levels to SARS-CoV-1 spike protein were very low (essentially negative) in the pre-pandemic healthy controls. However, SARS-CoV-1 spike-reactive antibodies increased significantly after SARS-CoV-2 infection. These increases were quite striking for IgG (p=0.0038) and also IgA (p=0.0084) and most likely represent cross-reactive antibodies directed to SARS-CoV-2 spike epitopes that are conserved between SARS-CoV-2 and SARS CoV-1^14^. These newly induced cross-reactive IgG antibodies generated after COVID-19 infection declined with an estimated half-life of 215 days (95%CI [168, 298]) (exponential decay model) (Figure 2). Taken together, these results show that people infected with SARS-CoV-2 may have also have some heightened immunity against the common human betacoronaviruses and more importantly against SARS-CoV-1.

### Durable neutralizing antibody responses to SARS-CoV-2 in infected patients

Neutralizing antibodies were measured with a live virus focus reduction neutralization test that uses a recombinant SARS-CoV-2 virus expressing the fluorescent reporter gene mNeonGreen (FRNT-mNG) (Figure 3A). During the first 250 days post-symptom onset, FRNT_50_ titers varied considerably between individuals and ranged from <20 to 3726 (Figure 3A). Of the 183 individuals for whom longitudinal neutralization titers were assayed, 140 (77%) had at least one timepoint with neutralization titers above the limit of detection (>20). Seventy-five percent (43/57) of COVID-19 patients generated serum neutralizing antibodies between 30 – 50 days after symptom onset, and similarly 72% (48/67) had measurable titers between 180 – 263 days after symptom onset. Using an exponential decay model, we evaluated the kinetics of neutralizing antibody titers after day 42 and estimated a half-life of 150 days (95%CI [124, 226]). However, similar to the spike reactive IgG binding antibodies, we hypothesized that the neutralizing antibody rate of decay may actually slow over time during the recovery period. To address this, we fit a power law to the data. The power law model fit significantly better than the exponential decay model (ΔAIC=9) and Δ estimated the half-life of neutralizing antibody responses at 120 days post-symptom onset to be 254 days (95%CI [183, 400]).

**Figure 3.**
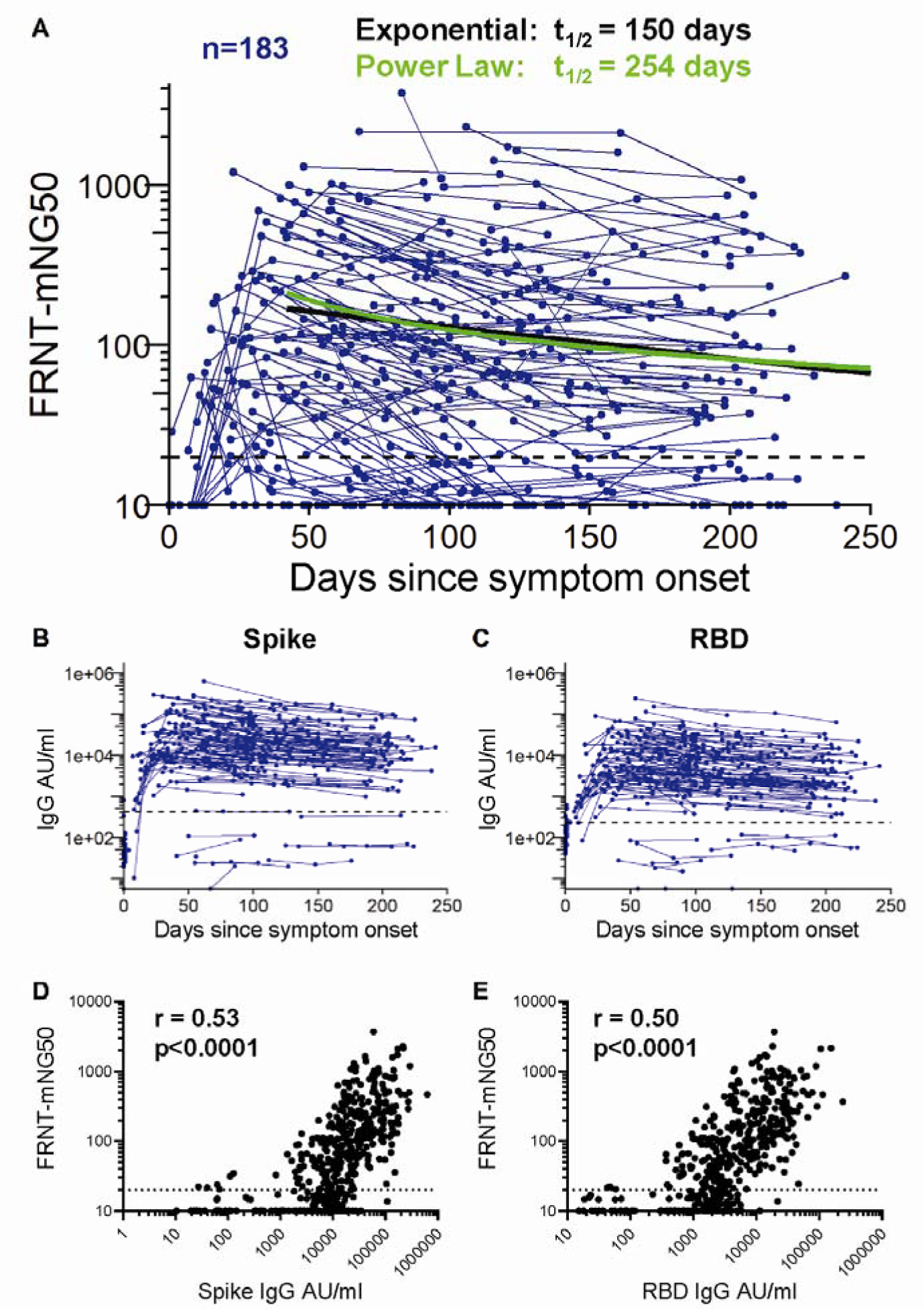
Neutralizing antibody responses to SARS-CoV-2. (A) *In vitro* serum neutralization antibody titers to SAR-CoV-2 were measured in duplicate by focus-reduction neutralization assay COVID-19 patients (n=183). The limit of detection is indicated with a dashed line at FRNT-mNG_50_ = 20. The half-life estimated by the exponential decay model (black) is 150 days, whereas the half-life estimated at day 120 using the power law model (green) is 254 days. IgG antibody titers reactive to SARS-CoV-2 spike (B) and RBD (C) of the matched 183 COVID-19 for whom neutralization titers were assessed. The geometric mean titer plus 3 standard deviations of pre-pandemic samples is indicated by a dashed line (B and C). SARS-CoV-2 spike (D) and RBD (E) reactive IgG levels correlated with neutralization titers at the matched time point (repeated measures correlation, p<0.0001). The limit of detection is indicated with a dashed line at FRNT-mNG_50_ = 20.

Next, we assessed the relationship between the levels of spike and RBD binding antibodies and SARS-CoV-2 neutralization. Figures 3B and C show the SARS-CoV-2 spike and RBD binding antibody response kinetics of the 183 participants for whom neutralization titers were assessed. These exhibited a wide range of antibody binding levels ranging from non-responders (n=11) who did not elicit antibody titers above those of pre-pandemic controls (defined as a COVID-19 patient titer below the mean pre-pandemic antibody titer plus 3 standard deviations, see dashed line on Figure 3B, C) to those with IgG levels >200,000 AU/ml. Spike and RBD binding IgG levels correlated significantly with the neutralization titers (Figure 3D, E; p<0.0001).

Taken together, our findings show that induction of neutralizing antibodies occurs in the majority of COVID-19 patients. These neutralizing antibodies can persist over the 8-9 month period following infection, and show a correlation with spike and RBD binding IgG.

### SARS-CoV-2 spike and RBD-specific memory B cells increase for several months after infection and then plateau over 8 months

Memory B cells (MBC) are an important component of humoral immunity and contribute to viral control by generating antibody responses upon re-exposure to the pathogen. We used full-length spike and RBD antigen probes to quantify the frequencies of SARS-CoV-2 spike-and RBD-specific MBC in longitudinal PBMC samples from 111 COVID-19 patients (Figure 4) and from 29 pre-pandemic controls (Figure S3A, B). Our flow cytometric gating strategy to identify SARS-CoV-2 specific MBC and classify them as IgG, IgM and IgA MBC isotypes is shown in Figure 4A.

**Figure 4.**
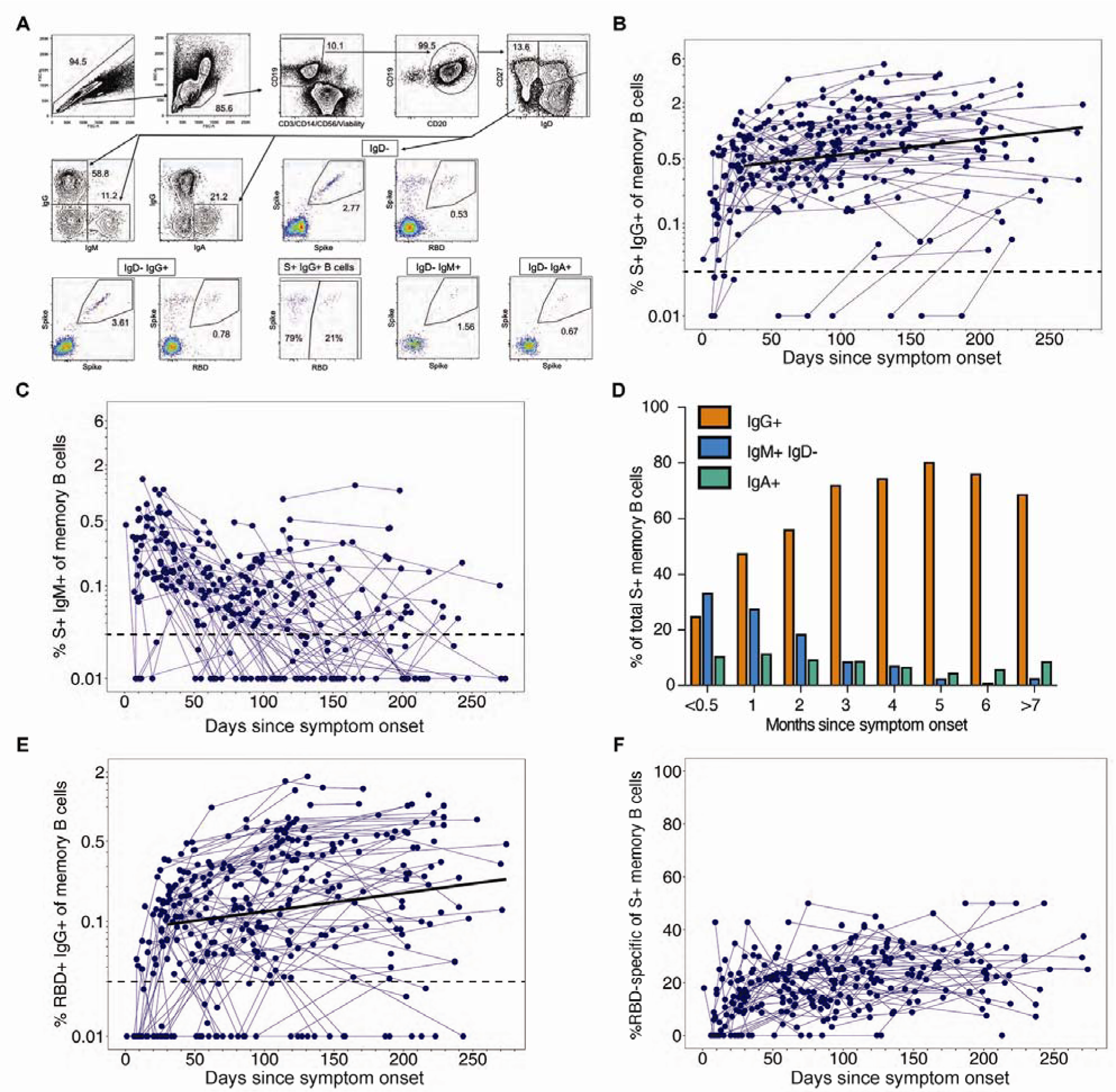
SARS-CoV-2 spike and RBD specific memory B cells. (A) Representative memory B cell gating strategy is shown for identification of SARS-CoV-2 spike and RBD-specific IgD-IgG+, IgD-IgM+ and IgD-IgA+ memory B cells in PBMCs from a SARS-CoV-2 convalescent participant. The frequency of spike+ (B) IgG+ and (C) IgM+ memory B cells out of memory B cells (IgD-CD19+ CD20+) is displayed over time from initial symptom onset among SARS-CoV-2 infected subjects (n=105 subjects; measured in singlet replicates). The dashed line indicates the limit of detection. The bold line represents the median fitted curve from a linear mixed effects model of post-day 30 responses. (D) The median percent of spike+ memory B cells expressing IgG, IgM or IgA isotypes was assessed at monthly intervals post-symptom onset. (E) The frequency of RBD+ IgG+ of memory B cells over time (n=141). (F) The proportion of S+ IgG+ memory B cells that are specific for the receptor binding domain are depicted over time.

Among the total MBC, the spike IgG+ MBCs were significantly increased in COVID-19 patients (n=111; Figure 4B) in comparison to pre-pandemic controls (n=29; Figure S3A) (median increase, 0.73% vs. 0.02%; p<0.0001). After a steep early expansion over the first 2-3 months, the spike IgG+ MBC persisted in COVID-19 patients with no decline out to 250 days post symptom onset. These findings (Figure 4B) are supported by a positive slope (0.004) from the model of the longitudinal spike IgG+ MBC responses after day 30 (95%CI [0.002, 0.006], p<0.001; Figures S4A, B).

The spike IgM+ MBC appeared within the first two weeks post-symptom onset and quickly declined (Figure 4C, D). The decay continued after day 30 (slope=-0.007, 95%CI [-0.010, -0.005], p<0.001). One month after symptom onset, 56% of spike MBC were IgG+, which increased to a peak of 80% at 5-6 months (Figure 4D). Circulating spike IgA+ MBC were also detectable in many subjects at low frequencies and without significant change over time (day 30 to 250: slope=0.000, 95%CI [-0.002, 0.002], p=0.91, Figure 4D).

Since the RBD contains the primary neutralizing epitopes on the spike, we also used an RBD-specific probe to characterize this subset of spike-specific memory B cells. Overall, approximately 20% of the spike IgG+ memory B cells targeted the RBD, which was consistent across subjects and time (Figure 4E, F). As expected, RBD+ IgM+ MBC emerged early in infection and subsequently switched to RBD+ IgG+ MBCs, which gradually increased during follow-up (day 30 to 250: slope=0.004, 95%CI [0.002, 0.005], p<0.001, Figure 4E). Thus, the maintenance of circulating spike-and RBD-specific IgG memory B cells suggests that these cells could be recruited for a rapid secondary response following re-exposure or vaccination.

### Induction of durable and polyfunctional virus specific memory CD4+ and CD8+ T cells in infected patients

CD4+ T cells are critical for generation of high affinity antibody responses and can also have anti-viral effects. In addition, they provide help for CD8+ T cell responses, which are vital for killing infected cells and mediating viral clearance. Thus, we next examined virus-specific CD4+ and CD8+ T cell responses longitudinally in COVID-19 patients and uninfected controls using a high-dimensional, multi-parameter *ex vivo* intracellular cytokine staining (ICS) assay. The assay is sensitive, precise and specific for detection of antigen-specific T cells expressing multiple cytokines and effector molecules following a short-term (6 hours) stimulation with peptide pools. Our lab developed and validated the assay, and we are currently using the method to quantitate Th1/Th2 CD4+ and CD8+ T cell responses in SARS-CoV-2 vaccine trials. Here, we assessed T cell responses to the SARS-CoV-2 structural (S, E, M and N) and accessory proteins (ORF 3a, 6, 7a, 7b, and 8) using overlapping peptide pools that span the sequences of these proteins.

Among COVID-19 patients, 89% (102/113) mounted CD4+ T cell responses (Figure 5A) recognizing at least one SARS-CoV-2 structural protein that was detectable at one or more visits. By contrast, SARS-CoV-2 specific CD4+ T cells were rarely detected in the uninfected control group using this assay (Figure S3C). Antigen-specific CD4+ T cells expanded over the first month after infection and then gradually declined over subsequent months. Their estimated half-life was 207 days (95%CI [104, 211]) as shown in Figure 5A, and these findings are supported by the individual CD4+ T cell response levels and slopes after day 30 (slope= -0.0033, 95%CI [-0.0017, -0.0066], p<0.0001) (Figure S4C, D). Of note, we observed a wide range in the total magnitude of responses, some reaching >1% of circulating CD4+ T cells, and an overall median frequency of 0.51% (Figures 5A and S5).

**Figure 5.**
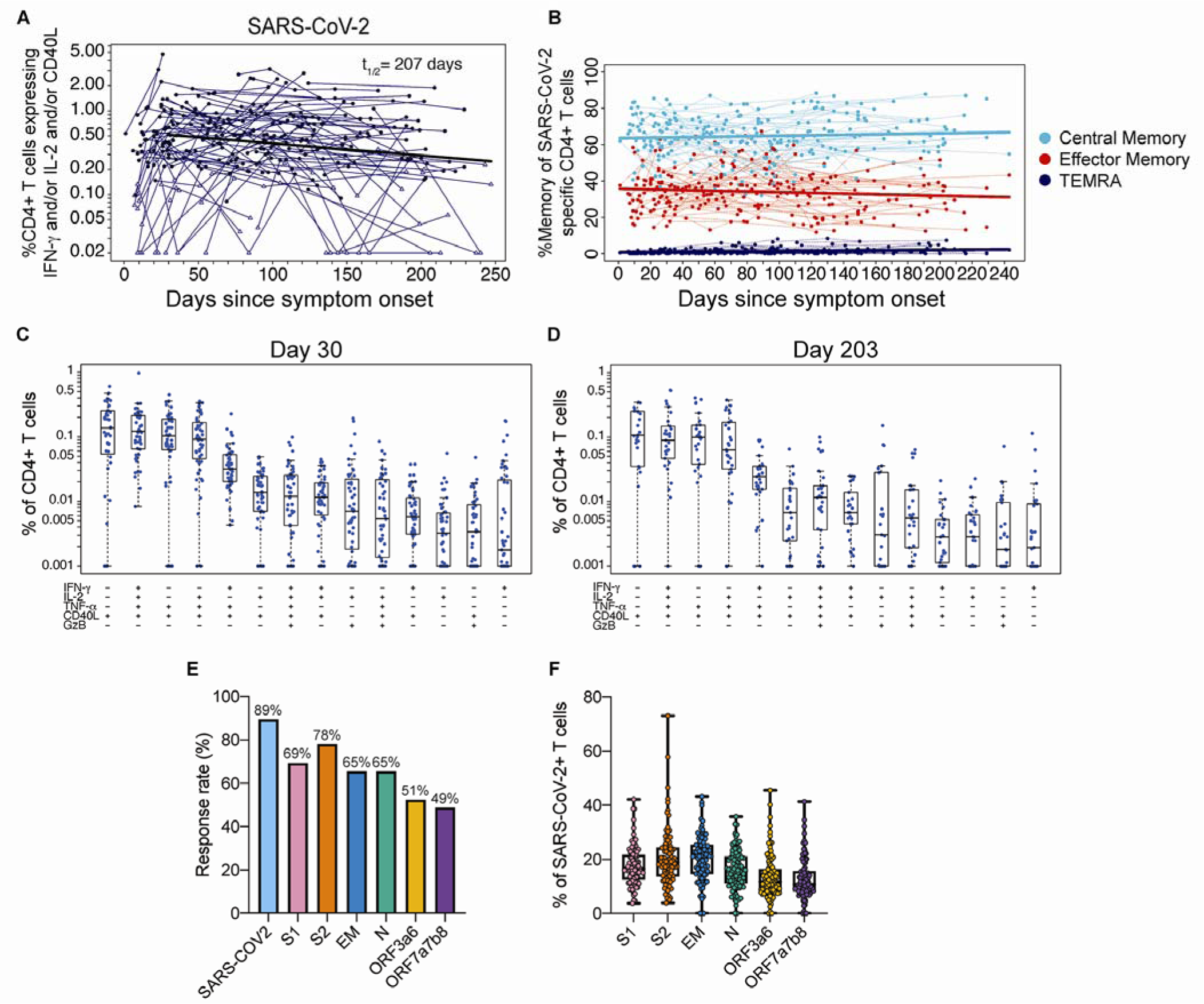
CD4+ T cell responses to SARS-CoV-2 antigens. (A) The sum of background-subtracted CD4+ T cells expressing *ex vivo* IFN-γ, IL-2 and/or CD40L to peptide pools spanning SARS-CoV-2 structural proteins: S1, S2, envelope (E), membrane (M), nucleocapsid (N), and the following ORFs: 3a, 3b, 6, 7a, 7b, and 8 (n=114; tested in singlets) for each individual/timepoint. Each sample that is ‘positive’ (by MIMOSA) for at least one SARS-CoV-2 antigen is indicated by a solid circle, whereas samples that are ‘negative’ for all of the SARS-CoV-2 antigens at that timepoint are indicated by open triangles. The bold line represents the median fitted curve from a nonlinear mixed effects model of post-day 30 responses among those with a positive response at >= 1 time point; t_1/2_ is the median half-life estimated from the median slope, with 95%CI [104, 411]. (B) The proportion of SARS-CoV-2-specific CD4+ T cells expressing a specific memory phenotype over time: central memory (CCR7+ CD45RA-), effector memory (CCR7-CD45RA-) or T_EMRA_ (CCR7-CD45RA+); restricted to ‘positive’ responders. Polyfunctionality of SARS-CoV-2-specific CD4+ T cells are shown at (C) 21-60 days since symptom onset (median, 30 days) and (D) >180 days median post symptom onset (median, 203 days). Percentages of cytokine-expressing CD4+ T cells are background subtracted and only subsets with detectable T cells are displayed. Data shown were restricted to ‘positive’ responders and a single data point per individual per time frame. All subsets were also evaluated for expression of IL-4, IL-5, IL-13, IL-17 and perforin, and were found to be negative. (E) Bar graphs indicate the proportion of COVID-19 convalescent patients who had a positive CD4+ T cell response to the individual SARS-CoV-2. peptide pool *ex vivo* stimulations. Some antigens were combined for stimulation as indicated. (F) For each subject with positive SARS-CoV-2-specific CD4+ T cells, the proportion of the total SARS-CoV-2 responding CD4+ T cells that are specific for each stimulation.

To better characterize the development of T cell memory in SARS-CoV-2 infection, we examined the differentiation profiles of virus-specific T cells longitudinally in COVID-19 patients. Based on CD45RA and CCR7 expression, SARS-CoV-2-specific CD4+ T cells were primarily central memory phenotype (CD45RA-CCR7+) and to a lesser extent effector memory (CCRA-CCR7-); this profile of the memory T cell subsets was very consistent between subjects and stable over time (Figure 5B). The antigen-specific CD4+ T cells were Th1-biased with a predominant CXCR3+CCR6-phenotype, andhighly polyfunctional, with simultaneous detection of antigen-specific CD154, IFN-γ, IL-2, TNF-α and less frequently granzyme B in the early expansion phase (21-60 days post symptom onset; median, 30 days) (Figure 5C). Interestingly, many of the virus-specific CD4+ T cells also exhibited this polyfunctionality at the memory time point (>180 days post symptom onset; median, 203 days) (Figure 5D). Circulating SARS-CoV-2-specific Th2 (IL-4, IL-5 and IL-13), Th17 (IL-17) or perforin-expressing subsets were not detected (Figure 5C and D).

Next, we examined the CD8+ T cell responses in COVID-19 patients and found that 69% generated CD8+ T cells recognizing at least one SARS-CoV-2 structural protein that were detectable at one or more visits (Figure 6A), in contrast to infrequent to rare, low-level antigen specific responses in the uninfected control donors (Figure S3D).

**Figure 6.**
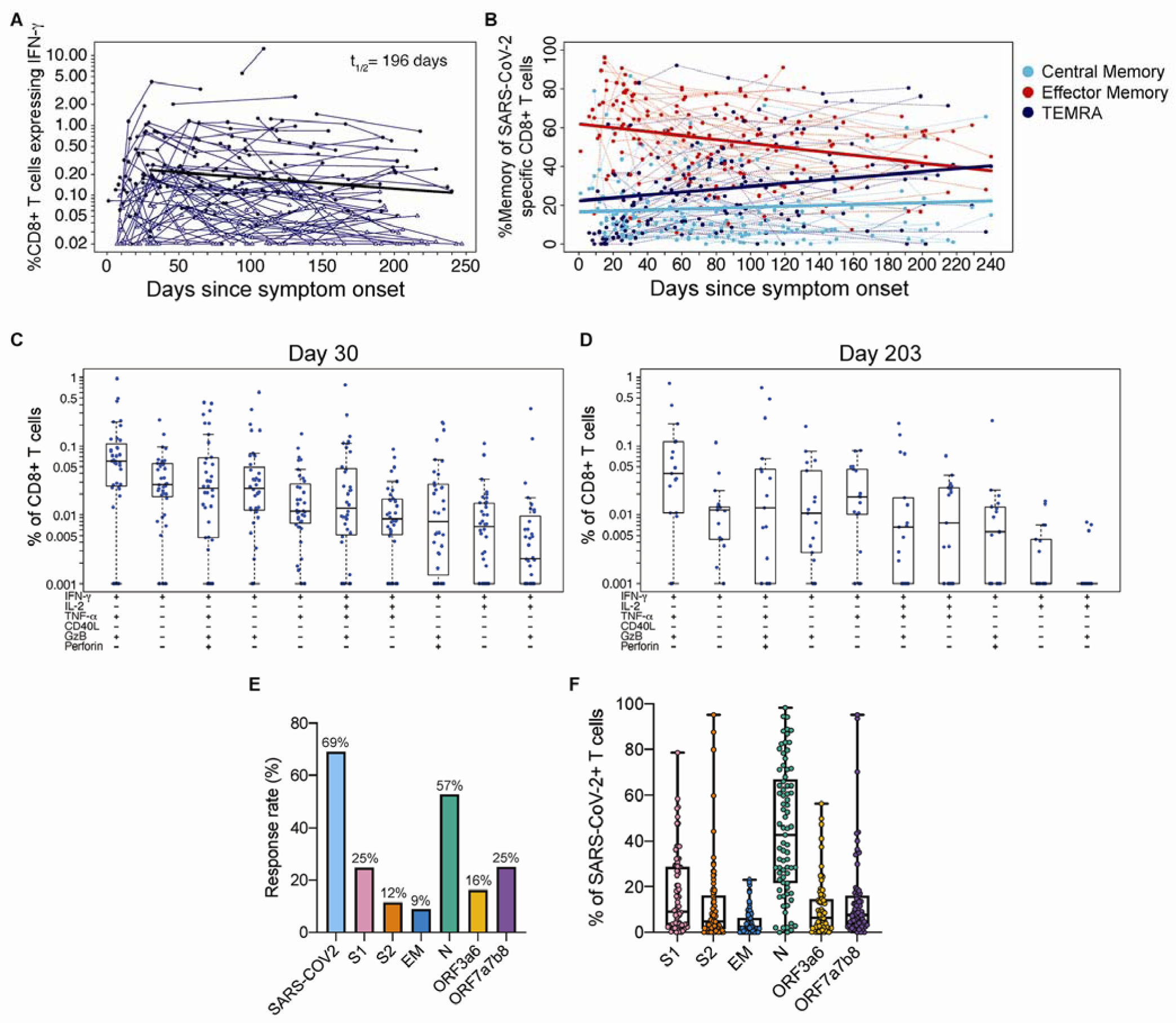
CD8+ T cell responses to SARS-COV-2 antigens. (A) The sum of background-subtracted CD8+ T cells expressing IFN-γ (with or without other cytokines), in response to peptide pools covering SARS-CoV-2 structural proteins: S1, S2, envelope (E), membrane (M), nucleocapsid (N), and the following ORFs: 3a, 3b, 6, 7a, 7b, and 8 (n=114; tested in singlets) for each individual/timepoint. Each sample that is ‘positive’ (MIMOSA) for at least 1 SARS-CoV-2 antigen is indicated by a solid circle, whereas samples that are ‘negative’ for all of the SARS-CoV-2 antigens at that timepoint are indicated by open triangles. The bold black line represents the median fitted curve from a nonlinear mixed effects model of post-day 30 responses among those with a positive response to the antigen(s) under consideration at ≥ 1 time point; t_1/2_ shown is the median half-life estimated from the median slope, with 95%CI [92, 417]. (B) The proportion of SARS-CoV-2-specific CD8+ T cells by memory phenotype over time: effector memory (EM; CCR7-CD45RA-), T_EMRA_ (CCR7-CD45RA+) and central memory (CM; CCR7+ CD45RA-). Analyses were restricted to ‘positive’ responders. Polyfunctionality of SARS-CoV-2-specific CD8 T cells at (C) 21-60 days post symptom onset (median, 30 days) and (D) >180 days median post symptom onset (median, 203 days). Percentages of cytokine expressing CD8+ T cells are background subtracted and only subsets with detectable T cells are displayed. Data shown were restricted to positive responders and a single data point per individual per time frame. All CD8+ T cell subsets were also evaluated for expression of IL-4, IL-5, IL-13, and IL-17 and were found to be negative. (E) The bar graphs indicate the proportion of COVID-19 convalescent patients who had a ‘positive’ CD8+ T cell response to the individual SARS-CoV-2 stimulations. (F) The fraction of the total SARS-CoV-2 responding CD8+ T cells per subject that are specific for each peptide pool.

Expansion of CD8+ T cells occurred over the first month and then frequencies gradually declined, with a half-life of 196 days (95%CI [92, 417]) and a negative estimated slope after 30 days of symptom onset (slope=-0.004, 95%CI [-0.002, -0.008], p<0.0001) (Figure 6A). The median frequency of SARS-CoV-2-specific CD8+ T cells was 0.2%, indicating a lower overall response magnitude than observed for CD4+ T cells.

However, like the CD4+ T cells, a wide range in magnitudes was observed with many SARS-CoV-2-specific CD8+ T cell frequencies above 1% and even up to 12% (Figure 6A).

A very different pattern of phenotypic changes were observed with virus-specific CD8+ T cells compared to what we saw with the CD4+ T cells (Figure 6B vs. Figure Figure 5B). In contrast to the dominance of the central memory subset with SARS-CoV-2-specific CD4+ T cells, the vast majority of the virus-specific CD8+ T cells showed an effector memory phenotype during the early phase of the response. However, this population of SARS-CoV-2-specific effector memory (CD45RA-CCR7-) contracted over time (slope=-0.904, p<0.0001; Figure 6B) and simultaneously there was an increase in the proportion of the TEMRA (CD45RA+CCR7-) subset of virus-specific CD8+ T cells (slope=0.075, p<0.0001; Figure 6B). A small but stable fraction of SARS-CoV-2-specific CD8+ T cells expressed a central memory phenotype (slope=0.024, p=ns; Figure 6B).

The SARS-CoV-2-specific CD8+ T cells were highly polyfunctional with the highest, TNF-α and granzyme B; other dominant magnitude populations secreting IFN-γ subsets also expressed IL-2 or perforin (Figure 6C, D). This polyfunctional profile was seen in the expansion phase (median 30 days; Figure 6C) and also at the later time points (>180 days post symptom onset; median 203 days; Figure 6D). It is important to note that this pattern of CD8+ T cell differentiation has been described in detail after vaccination in humans with the live attenuated yellow fever virus vaccine (YFV-17D).^15^ This YFV-17D vaccine generates long-lived and functional virus specific memory CD8+ T cells that persist in humans for decades.^15, 16^ That the CD8+ T cell differentiation program after COVID-19 infection resembles what is seen after YFV infection of human suggests that COVID-19 patients may also generate long-lived CD8+ T cell memory.

### CD4+ and CD8+ cells target different SARS-CoV-2 antigen specificities

The majority of COVID-19 patients generated CD4+ T cells that recognized most SARS-CoV-2 viral structural and accessory proteins, with the highest percentage responding to S2 (78%) and S1 (69%) (Figure 5E, F). Among the COVID-19 subjects with positive responses, the proportion of SARS-CoV-2-specific CD4+ T cells reacting to each peptide pool was evenly distributed (Figure 5F). Thus, CD4+ T cells equally targeted multiple SARS-CoV-2 proteins.

In contrast to the results seen with CD4+ T cells, SARS-CoV-2-specific CD8+ T cells showed preferential recognition of the nucleocapsid protein. The dominant CD8+ T cell response rate was directed to the nucleocapsid (57%), followed by ORFs 7a, 7b, and/or 8 (25%), S1 (25%), ORFs 3a and/or 6 (16%), S2 (12%) and E and/or M (9%) (Figure 6E). Also, among the COVID-19 patients with CD8+ T cell responses, there was a bias with the largest percentage (median, 43%) reacting to the nucleoprocapsid protein (Figure 6F). While SARS-CoV-2 CD8+ T cell responses rates were much lower in uninfected controls, when present in a few control donors with lower frequencies, these were also targeted to the nucleocapsid protein (Figure S3D). A likely explanation for these findings is that in SARS-CoV-2 infection, antigen-presenting cells *in vivo* may display a higher proportion of peptides derived from the nucleocapsid protein and hence more nucleocapsid-specific CD8+ T cells are generated during infection. This has interesting implications suggesting that nucleocapsid-specific CD8+ T cells might be more efficient in recognizing virally infected cells.

### Age and disease severity are significantly associated with magnitude of SARS-CoV-2 immune responses

We evaluated whether COVID-19 patient age, disease severity, or gender could account in part for the heterogeneity observed among the SARS-CoV-2-specific immune responses as estimated from the individual models (post day 30 for cellular and post day 42 for antibody responses). We observed that age was significantly associated with higher immune responses to SARS-CoV-2, independently of any covariation with disease severity (Figure 7A). Neutralizing antibody titers and IgG antibody responses to nucleocapsid increased 1.35-fold and 1.25-fold, respectively, with each decade of age and the same disease severity (95%Cis [1.19, 1.54] and [1.08, 1.43], p values<0.003). Similarly, increased age positively correlated with increased frequencies of spike and RBD-specific IgG+ memory B cells, with 1.19 to 1.24-fold higher responses per decade of age (p values<0.02; Figure 7A), accounting for disease severity. Increased age also correlated with higher SARS-CoV-2 and S1-specific CD4+ T cell responses (1.16-1.20-fold increase by decade of age, p values<0.02) and N-specific CD8+ T cell responses (1.24-fold increase by decade of age, p=0.039) accounting for disease severity (Figure 7A).

**Figure 7.**
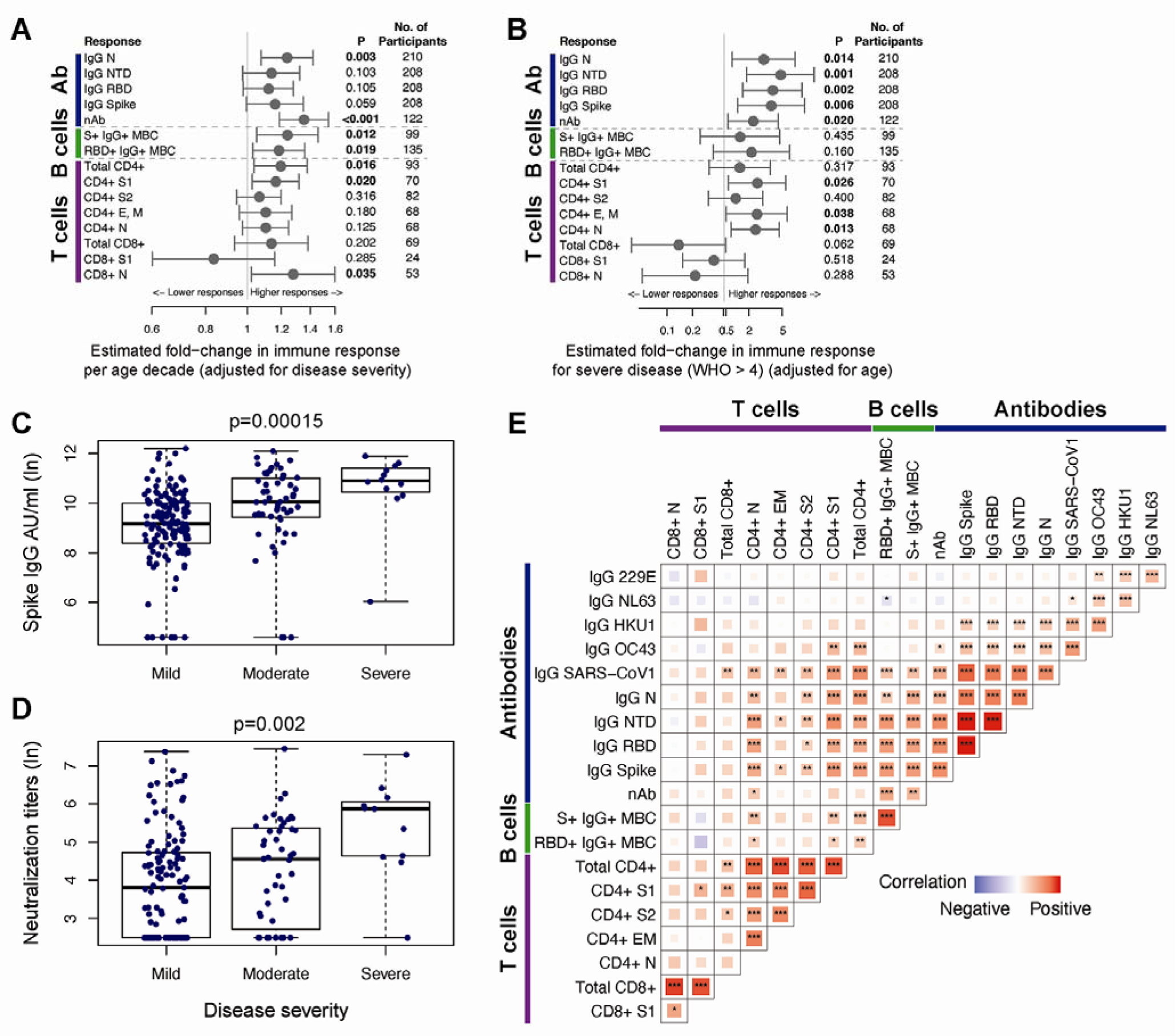
Correlations between SARS-CoV-2-specific immune responses and assessment of covariates. (A) The forest plot depicts the estimated fold-change in the level of each immune response per decade of age, with 95% Wald-based CIs and p-values. (B) The forest plot shows the estimated fold-change in the level of each immune response for severe (WHO score > 4) vs. non-severe (WHO score 4) disease, with 95% Wald-based CIs and p-values. S1 CD8+ T cell responses compared moderate-severe (WHO score > 2) to mild (WHO score 2) disease as there were no participants with severe disease with at least one positive S1 CD8+ T cell response post-day 30. Estimates in (A) and (B) are from mixed effects models of post-day 30 (B and T cell responses) or post-day 42 (antibody responses) among responders that account for fixed effects of age and disease severity on the level of immune response. Univariate assessment of disease severity on the magnitude of (C) spike IgG antibodies and (D) SARS-CoV-2 neutralizing antibodies at day 120 is shown for mild (WHO score: 0-2), moderate (WHO score: 3-4), and severe disease (WHO score: 5+); p-values from one-way ANOVA. (E) The heatmap shows Spearman correlations between critical SARS-CoV-2 memory immune responses (day 30 B and T cell responses and day 180 antibody responses) with significance levels: *p<0.05, **p<0.01, ***p<0.001. The tile size and color intensity correspond to the absolute value of the Spearman rank correlation coefficient, with red or blue indicating a positive or negative correlation, respectively. Day 30, 42 and 180 immune responses were estimated from mixed effects models of the longitudinal SARS-CoV-2 binding antibodies, SARS-CoV-2 neutralizing antibodies, CD4+ and CD8+ T cell responses, and B cell responses.

Since the cohort included primarily persons with mild to moderate COVID-19, we had limited ability to assess the relationship of severe disease and SARS-CoV-2 immune responses, especially among the cellular responses. However, we found that after accounting for age, severe disease (WHO score >4) was associated with higher IgG antibodies to nucleocapsid, spike, RBD, and NTD (Figure 7B, C), and SARS-CoV-2 neutralization titers (Figure 7D). Severe disease was also associated with 2.30 to 2.46-fold higher S1, E and/or M, and nucleocapsid-specific CD4+ T cells (all p values<0.05; Figure 7B). We found no significant relationships between gender and the immune responses evaluated, apart from 1.66-fold higher IgG NTD responses antibodies among males compared to females, after accounting for age and disease severity (95%CI [1.08, 2.55], p=0.022). In all, our analyses suggest that there are synergistic, but also independent mechanisms driving higher adaptive immune responses in COVID-19 patients who are older and/or who experienced more severe disease.

### Early SARS-CoV-2 B and T cell responses correlated with durable spike and RBD IgG antibody binding and neutralization titers

We assessed correlations between SARS-CoV-2-specific immune responses using the individual-level models to interpolate the magnitude of responses for each COVID-19 patient at early (day 30) or later (day 180) convalescent time points (Figure 7E). We found that durable serum neutralization titers correlated with the magnitude of IgG+ binding antibodies to spike, NTD and RBD at day 180 each (day 180; Spearman R=0.62, 0.61, and 0.61, respectively; all p values<0.0001). Similarly, the frequency of RBD+ IgG+ memory B cells at day 30 correlated with the maintenance of RBD+ IgG antibodies (day 180; Spearman R=0.53, p<0.0001) and neutralization antibody titers (day 180; Spearman R=0.48, p<0.0001). We also observed that the magnitude of S1-specific CD4+ T cells at day 30 correlated with durable IgG antibodies against spike (day 180; Spearman R=0.56, p<0.0001), NTD (Spearman R=0.62, p<0.0001), and RBD (Spearman R=0.47, p=0.0002) (Figure 7E). These findings are consistent with early SARS-CoV-2 memory B cells and CD4+ T cells supporting the generation of durable antibody responses.

## DISCUSSION

Establishing immune memory is essential in the defense against SARS-CoV-2 infection. To end the COVID-19 pandemic, it is critical to know how long immunity against SARS-CoV-2 will persist after infection and whether it will be sufficient to prevent new infections and severe disease in years to come. Identifying in depth the adaptive immune components leading to recovery and modeling the trends of each response was enabled by the longitudinal sampling of a large number of COVID-19 patients. Here, we show that most convalescent COVID-19 patients mount durable antibody, B and T cells specific for SARS-CoV-2 up to 250 days, and the kinetics of these responses provide an early indication for a favorable course ahead to achieve long-lived immunity. Because the cohort will be followed for 2-3 more years, we can build on these results to define the progression to long-lived immunity against this novel human coronavirus, which can guide rational responses when future outbreaks occur.

The hallmark of the initial immune defense against SARS-CoV-2 is the emergence of antibodies recognizing the SARS-CoV-2 spike protein, including the RBD and NTD components of the S1 subunit, during the early phase of viral replication. These antibodies are likely secreted from plasmablasts rapidly generated from B cells that are activated upon their first encounter with the pathogen spike antigen. The brisk rise over the first month of infection, followed by a fast decline of the circulating spike IgG and IgA antibodies, is a consistent finding and likely explained by the disappearance of the short-lived plasmablasts. These events occur even sooner for the spike IgM and nucleocapsid antibodies.

Some antibodies that bind to specific epitopes on the spike RBD and NTD can block SARS-CoV-2 infection of respiratory epithelial cells by inhibiting the interactions of the viral spike with the ACE2 receptor.^17–20^ Thus, as expected, the early rise and decline of antibodies neutralizing live SARS-CoV-2 were similar to the kinetics of antibodies binding the spike and RBD protein. The striking finding is the bi-phasic curve of the spike-specific binding and neutralizing antibody responses when analyzed with the power law model, which provides a better fit for the antibody kinetics after the peak response.^21^ This bi-phasic decline accords with other recently published observations on SARS-CoV-2 serological kinetics.^22, 23^ With sampling data extended to 250 days, we were able to detect a slowing of the decay of these functional antibodies toward a plateau level, suggestive of the generation of longer-lived plasma cells, and durable antibody responses. The importance of these observations is that following recovery, neutralizing antibodies may persist, albeit at low levels, and may act as the first line of defense against future encounters of SARS-CoV-2 and possibly related human coronaviruses.

Another interesting finding of this investigation is the remarkably stable antibody responses among the pre-pandemic and COVID-19 patients to the common human coronaviruses that are acquired in children and adults. These data are most consistent with the generation of long-lived plasma cells and refute the current notion that these antibody responses to human coronaviruses are short-lived. Moreover, the COVID-19 patients mounted increased IgG antibody responses to SARS-CoV-1, a related pathogen that none likely had experienced previous exposure. This finding is consistent with the booster response of SARS-CoV-1 neutralizing antibodies that we recently observed following SARS-CoV-2 mRNA vaccination.^3, 24^ Taken together, these results may have implications for a broader strategy for vaccines targeting multiple betacoronaviruses.

The durable antibody responses in the COVID-19 recovery period are further substantiated by the ongoing rise in both the spike and RBD memory B cell responses after over 3-5 months before entering a plateau phase over 6-8 months. Persistence of RBD memory B cell has been noted.^25–27^ We presume this may be explained by sustained production of memory B cells in germinal centers of lymph nodes draining the respiratory tract in the early months, followed by the memory B cell redistribution into the circulation as the germinal centers begin to recede. Thus, the induction and maintenance of memory B cells and, over time, long-lived plasma cells, will continue to furnish higher affinity antibodies if re-exposures occur.

In contrast to spike memory B cell kinetics, SARS-CoV-2-specific CD4+ and CD8+ memory T cells each peak early, within the first month, but then slowly decline over the next 6-7 months. Central memory Th1-type CD4+ T cells dominate throughout the early infection and recovery period. However, the CD8+ T cells exhibit a predominant effector memory phenotype early that transitions to those effector memory cells re-expressing CD45RA, maintaining expression of antiviral cytokines and effector functions which have been shown to provide protective immunity against other viral pathogens.

We also provide clear evidence that the CD4+ T cells mount a broader antigen-specific response across the structural and accessory gene products, whereas the CD8+ T cells are predominantly nucleocapsid specific and spike-specific responses are substantially lower in frequency.

Our study demonstrates the considerable immune heterogeneity in the generation of potentially protective response against SARS-CoV-2, and by focusing on the dynamics and maintenance of B and T cell memory responses, we were able to identify features of these early cellular responses that can forecast the durability of a potentially effective antibody response. The ability to mount higher frequencies of RBD-specific memory IgG+ B cells early in infection was the best indicator for a durable RBD-specific IgG antibody and neutralizing antibody response. In addition, higher frequency CD4+ T cells were associated with stronger spike IgG and neutralizing antibody responses.

However, the induction and peak response of SARS-CoV-2-specific CD8+ T cells occurs independently to these antibody responses. Interestingly, while it has been widely reported that age correlates with COVID-19 disease severity, we found that age and disease severity were independent co-variates associated with the magnitude of both SARS-CoV-2-specific CD4+ T cell and humoral SARS-CoV-2 immunity, but not with the magnitude of CD8+ T cell responses. In the case of T cells, whether the T cell differences are related to the frequencies or specificities of pre-existing coronavirus CD4+ and CD8+ T cell immunity will require additional future analysis.

The COVID-19 pandemic remains a global public health threat after one year of overwhelming disruption and loss. Overcoming the challenges to end the pandemic is accentuated by the recognition that SARS-CoV-2 can undergo rapid antigenic variation that may lower vaccine effectiveness in preventing new cases and progression to severe disease.^24, 28, 29^ Our findings show that most COVID-19 patients induce a wide-ranging immune defense against SARS-CoV-2 infection, encompassing antibodies and memory B cells recognizing both the RBD and other regions of the spike, broadly-specific and polyfunctional CD4+ T cells, and polyfunctional CD8+ T cells. The immune response to natural infection is likely to provide some degree of protective immunity even against SARS-CoV-2 variants because the CD4+ and CD8+ T cell epitopes will likely be conserved. Thus, vaccine induction of CD8+ T cells to more conserved antigens such as the nucleocapsid, rather than just to SARS-CoV-2 spike antigens, may add benefit to more rapid containment of infection as SARS-CoV-2 variants overtake the prevailing strains.

### Limitations of study

Our study evaluates COVID-19 patients only up to 8 months and requires models to estimate immune response half-lives thereafter. Because our longitudinal study will extend beyond two years, we can corroborate our models with subsequent experimental data on the persistence of immune memory. Our study population was primarily outpatients with mild to moderate COVID-19 and thus we were unable to evaluate immune memory in those with the extreme presentations, both asymptomatic and severe COVID-19. However, mild-moderate illness accounts for nearly all cases of COVID-19 and highlights the relevance of our findings over time.

## Data Availability

All data is provided in the manuscript.

## ACKNOWLEDGMENTS

First, we thank the participants for volunteering their time and effort to participate in this study. We thank Children’s Healthcare of Atlanta, the Georgia Research Alliance, and the Donaldson Trust for their support. The Emory Children’s Center-Vaccine Research Clinic also thanks Laila Hussaini, Ashley Tippett, Amy Muchinsky, and Sydney Biccum for their assistance with this study. At the Hope clinic of Emory Vaccine Center, we thank Rebecca Fineman, Dumingu Nipuni Gomes, Ellie Butler, Michelle Wiles for their assistance with the study. At the Fred Hutchinson Cancer Research Center, we thank Roland Strong for providing recombinant SARS-CoV-2 hexapro spike (S6P) and Leo Stamatatos for providing RBD protein. We also thank Rebecca Putnam, Todd Haight, Kim Louis, Ro Yoon, Carol Marty, Daryl Morris, Xiaoling Song, Mark Majeres, Joe Abbott, Omolara Akingba, Josh Donahue, Tu Anh Nguyen, Katharine Schwedhelm, Carly Sprague, and Terri Stewart for their vital assistance with this study.

## AUTHOR CONTRIBUTIONS

M.J.M and R. Ahmed conceived the study. M.J.M., S.E., J.C., E.J.A., A.K.M, N.R. and J.O.K. established the cohort and recruited the participants. S.L.L., M.P.L., C.W.D., M.P.G., S.G., K.A.S., G.M., C.N., V.V.E., L.L. and D.S.S. conducted serological assays and related analyses. H.A., V.I.Z., B.P. and Z.M. conducted formal statistical analyses and modeling. K.W.C., R.W. and L.E.N. planned, performed and analyzed antigen-specific B cell flow cytometry. S.C.D., K.W.C. and S.F. conceived, supervised, performed, and analyzed T cell experiments. V.E.E, K.F., and L.L. performed FRNT assays. K.W.C., S.L.L. and Z.M. drafted the original manuscript; M.J.M., M.S.S. and R.Ahmed edited the manuscript. All authors read and approved the manuscript. M.J.M., R.A, J.W. and M.S.S. secured funds and supervised the project.

## Funding Acknowledgments

The research reported in this publication was supported in part by COVID supplements from the National Institute of Allergy and Infectious Diseases and the Office of the Director of the National Institutes of Health under award numbers UM1AI068618-14S1 and UM1AI069481-14S1 (MJM); UM1A057266-S1, U19AI057266-17S1, 1U54CA260563, and U19AI090023 (R. Ahmed); ORIP/OD P51OD011132 (MSS); and T32AI074492 (LEN). This work was also supported by grants from the Oliver S. and Jennie R. Donaldson Charitable Trust (R. Ahmed); Paul G. Allen Family Foundation Award #12931 (MJM); Seattle COVID-19 Cohort Study (Fred Hutchinson Cancer Research Center, MJM); the Joel D. Meyers Endowed Chair (MJM); An Emory EVPHA Synergy Fund award (MSS and JW); COVID-Catalyst-I^3^ Funds from the Woodruff Health Sciences Center (MSS); the Center for Childhood Infections and Vaccines (MSS and JW); Children’s Healthcare of Atlanta (MSS and JW), a Woodruff Health Sciences Center 2020 COVID-19 CURE Award (MSS) and the Vital Projects/Proteus funds. The content is solely the responsibility of the authors and does not necessarily represent the official views of the funders.

## RESOURCE AVAILABILITY

### Lead contact

Further information and requests for resources and reagents should be directed to and will be fulfilled by the lead contacts: M. Juliana McElrath; and Rafi Ahmed.

### Materials Availability

This study did not generate new unique reagents.

### Data and code availability

All data and code reported in this paper will be shared by the lead contact upon request.

## EXPERIMENTAL MODEL AND SUBJECT DETAILS

### Study Populations

Two longitudinal COVID-19 cohort studies at Fred Hutchinson Cancer Research Center (Seattle, Washington) and Emory University (Atlanta, Georgia) began after receiving institutional review board approvals (IRB 10440, IRB 00001080 and IRB00022371). Adults ≥18 years were enrolled who met eligibility criteria for SARS-≥ CoV-2 infection and provided informed consent. Study participants provided medical history of co-morbidities, presentation of SARS-CoV-2 infection onset and disease course, and peripheral blood at initial and follow up visits for analysis of serum antibody and cellular immune responses. Additional longitudinal archived sera and PBMC from pre-pandemic study populations from Emory and Seattle served as controls for the immune assays.

The Atlanta study population included adult volunteers over the age of 18 who were diagnosed with COVID-19 by a commercially available SARS CoV-2 PCR assay, rapid antigen test, or clinical syndrome only (later confirmed with serology) due to limited SARS-CoV-2 testing during the early period of the pandemic. Ambulatory participants were recruited through local advertisements, internet-based avenues (such as social media, listserves), COVID-19 testing sites, and primary care clinics. Hospitalized patients were identified through SARS-CoV-2 testing. Informed consent was obtained from all participants prior to conduct of study procedures. Initial acute peripheral blood samples were collected from hospitalized patients at the time of enrollment.

Convalescent samples from hospitalized patients were collected when the patients were able to return for a visit to the clinical research site at the next study visit. Serial peripheral blood samples were collected starting at about 30 days after the onset of COVID-19 symptoms and/or after PCR positivity for SARS-CoV-2. Thereafter, samples were collected at 3, 6, and 9 months. The study is ongoing with expected completion of sample collection from participants in February 2023. Participants were excluded if they were immunocompromised, HIV positive, had active hepatitis B or C virus infection, used immunosuppressive drugs for 2 weeks or more in the preceding 3 months, received blood products or immune globulin 42 days prior to enrollment, received convalescent COVID-19 plasma, or were pregnant or breast feeding. We report on 110 participants to date, of which 73% were diagnosed by SARS-CoV-2 PCR, the remaining were diagnosed by rapid antigen test or serology. Demographic features of the participants are as follows: median age was 48; 45% were male; the majority (80%) were white, 11% Black/African American, 6% Asian, and 8% were Hispanic/Latinx ethnicity. The most frequent co-morbid conditions were hypertension, obesity, heart disease and diabetes mellitus. The most frequent COVID-19 symptoms were myalgia/fatigue, fever, cough, headache, loss of smell and taste (Table S1).

Hospitalized patients were older, with a median age of 56; a higher percentage were Black/African American (27%); and 100% had fever.

Longitudinal pre-pandemic sera samples from Emory were collected from individuals participating in a yellow fever vaccine study from 2014-2016 or an influenza vaccine study from 2015-2018^15, 30^. Data were included for analysis of binding antibody responses and are presented as days post-irrelevant (yellow fever) vaccination.

The Seattle COVID-19 Cohort study participants were recruited from the Seattle metropolitan area by social media advertisements, partnership with the local emergency medical service and by word of mouth. Study participants were screened and enrolled by the Seattle Vaccine Trials Unit staff. Eligibility criteria included adults at risk for SARS-CoV-2 infection or those diagnosed with COVID-19 by a commercially available SARS-CoV-2 PCR assay or blood antibody test and willing to have at least four blood draws collected over one year. Exclusion criteria included pregnancy and inability to donate blood.

Informed electronic consent was obtained from all Seattle participants during a screening phone call with study clinical staff. Interested participants were screened, consented and medical history and COVID-19 illness onset date and symptoms collected. Participants undiagnosed with COVID-19 had a nasopharyngeal (NP) swab collected and tested for SARS-CoV-2 via an FDA-approved PCR test and blood was collected for SARS-CoV-2 antibody (Abbott) and study assays. Those with either a positive PCR or antibody test were asked to return for future blood draws. Those who tested negative were asked to return as controls for the positive cohort and in case they tested positive in the future. Participants with a positive test prior to study enrollment or those diagnosed in study were asked to provide blood donation at approximately 7 days, 2 weeks, 1, 2, 3, 4, 6, 9-and 12-months post symptom onset. After completing one year of study, participants will be given the option of continuing the longitudinal study for up to two or more years. At each study visit, participant symptoms and medical history is updated. Those with COVID-19 symptoms after enrollment in all groups are offered a nasopharyngeal swab PCR SARS-CoV-2 test.

As of October 2020, 805 individuals have contacted the Seattle COVID-19 cohort study and 425 have enrolled. This includes 281 negative and 144 SARS-CoV-2 positive participants. Reasons for not enrolling include lack of interest, not meeting the eligibility criteria, inability to travel to blood draw location and inability to collect study blood. No participants have terminated from the study. Study enrollment and follow-up remains ongoing. Samples from SARS-CoV-2 negative subjects were included in B and T cell assays as ‘contemporaneous’ negative controls.

Peripheral blood mononuclear cells (PBMC) were obtained from HIV-1 seronegative donors who were recruited at the Seattle Vaccine Trials Unit before 2019 as part of the study “Establishing Immunologic Assays for Determining HIV-1 Prevention and Control”. All participants signed informed consent, and the Fred Hutchinson Cancer Research

Center IRB (Seattle, WA, USA) institutional human subjects review committee approved the protocol prior to study initiation. Pre-pandemic samples from this cohort were used as assay controls in B and T cell assays.

## METHOD DETAILS

### PBMC processing

PBMC for cellular assays were isolated by density centrifugation and cryopreserved from ACD-anticoagulated whole blood within eight hours of venipuncture, as described previously ^31^. Sera were also processed and cryopreserved within 4 hours after collection.

### Antibody binding assay

Antibody binding titers were measured using a multiplex plate coated with the SARS-CoV-2 spike, SARS-CoV-2 spike receptor binding domain, SARS-CoV-2 spike N terminal domain, SARS-CoV-2 nucleocapsid, SARS-CoV-1 spike, 229E spike, NL63 spike, HKU1 spike, and OC43 spike proteins (Mesoscale Discovery). Plates were blocked with 150μl/well with 5% bovine serum albumin in phosphate buffered saline μ (PBS) and shaken at 700 RPM at room temperature for at least 30 min. Plates were washed 3 times with 150μl/well 0.05% Tween-20 in PBS. Serum and plasma samples μ were added to the plate at dilutions between 1:500 and 1:50,000 and shaken at 700 RPM at room temperature for 2 hr. Following a wash, plates were incubated with 50ul/well of Sulfo-Tag anti-human IgG, IgA, or IgM detection antibody and shaken at 700RPM at room temperature for 1 hr. After a subsequent wash, 150μl/well of MSD μ GOLD read buffer was added to the plate and plates were immediately read on the MSD instrument to measure light intensity. Antibody levels are reported as arbitrary units/ml (AU/ml) based on normalization to a standard curve.

### Viruses and cells

VeroE6 cells were obtained from ATCC (clone E6, ATCC, #CRL-1586) and cultured in complete DMEM medium consisting of 1× DMEM (VWR, #45000-304), 10% FBS, 25mM HEPES Buffer (Corning Cellgro), 2mM L-glutamine, 1mM sodium pyruvate, 1× Non-essential Amino Acids, and 1× antibiotics. The infectious clone SARS-CoV-2 (icSARS-CoV-2-mNG), derived from the 2019-nCoV/USA_WA1/2020 strain, was propagated in VeroE6 cells (ATCC) and sequenced ^32^.

### Focus reduction neutralization test

Neutralization assays with SARS-CoV-2 virus were performed as previously described ^32–34^. Plasma/serum were serially diluted (three-fold) in serum-free Dulbecco’s modified Eagle’s medium (DMEM) in duplicate wells and incubated with 100–200 FFU infectious clone derived SARS-CoV-2-mNG virus at 37°C for 1 hr ^32^. The antibody-virus mixture was added to VeroE6 cell (C1008, ATCC, #CRL-1586) monolayers seeded in 96-well blackout plates and incubated at 37°C for 1 hr. Post-incubation, the inoculum was removed and replaced with pre-warmed complete DMEM containing 0.85% hr. After 24 hr, methylcellulose methylcellulose. Plates were incubated at 37°C for 2443 overlay was removed, cells were washed twice with PBS and fixed with 2% paraformaldehyde in PBS for 30 min at room temperature. Following fixation, plates were washed twice with PBS and foci were visualized on a fluorescence ELISPOT reader (CTL ImmunoSpot S6 Universal Analyzer) and enumerated using Viridot ^35^. The neutralization titers were calculated as follows: 1 - (ratio of the mean number of foci in the presence of sera and foci at the highest dilution of respective sera sample). Each specimen was tested in two independent assays performed at different times. The FRNT-mNG_50_ titers were interpolated using a 4-parameter nonlinear regression in GraphPad Prism 8.4.3. Samples with an FRNT-mNG_50_ value that was below the limit of detection were plotted at 20.

### Spike and RBD memory B cell flow cytometry assays

Fluorescent SARS-CoV-2-specific S6P^36^ (provided by Roland Strong, Fred Hutchinson Cancer Research Center, Seattle, WA) and RBD (provided by Leonidas Stamatatos, Fred Hutchinson Cancer Research Center, Seattle, WA) probes were made by combining biotinylated protein with fluorescently labeled streptavidin (SA). The S6P probes were made at a ratio of 1:1 molar ratio of trimer to SA. Two S6P probes, one labeled with AlexaFluor488 (Invitrogen), one labeled with AlexaFluor647 (Invitrogen), were used in this panel in order to increase specificity of the detection of SARS-CoV-2-specific B cells. The RBD probe was prepared at a 4:1 molar ratio of RBD monomers to SA, labeled with R-phycoerythrin (Invitrogen). Cryopreserved PBMCs from SARS-CoV-2-convalescent participants and a pre-pandemic SARS-CoV-2-naïve donor were thawed at 37°C and stained for SARS-CoV-2-specific memory B cells as described previously^19^ with a panel of fluorescently-labeled antibodies (see Key Resource Table). Cells were stained first with the viability stain (Invitrogen) in PBS for 15 min at 4°C. Cells were then washed with 2% FBS/PBS and stained with a cocktail of the three probes for 30 min at 4°C. The probe cocktail was washed off with 2% FBS/PBS and the samples were stained with the remaining antibody panel and incubated for 25 min at 4°C. The cells were washed two times and resuspended in 1% paraformaldehyde/1× PBS for collection on a LSR II or FACSymphony flow cytometer (BD Biosciences). Data was analyzed in Flow Jo version 9.9.4.

### Intracellular cytokine staining (ICS) assay

Flow cytometry was used to examine SARS-CoV-2-specific CD4+ and CD8+ T-cell responses using a validated ICS assay. The assay was similar to a published report ^5, 37, 38^ and the details of the staining panel are included in the Key Resource Table. Peptide pools covering the structural proteins of SARS-CoV-2 were used for the six-hour stimulation. Peptides matching the SARS-CoV-2 spike sequence (316 peptides, plus 4 peptides covering the G614 variant) were synthesized as 15 amino acids long with 11 amino acids overlap and pooled in 2 pools (S1 and S2) for testing (BioSynthesis). All other peptides were 13 amino acids overlapping by 11 amino acids and were synthesized by GenScript. The peptides covering the envelope (E), membrane (M) and nucleocapsid (N) were initially combined into one peptide pool, but the majority of the assays were performed using a separate pool for N and one that combined only E and M. Several of the open reading frame (ORF) peptides were combined into two pools: ORF 3a and 6, and ORF 7a, 7b and 8. All peptide pools were used at a final concentration of 1 μg/ml for each peptide. As a negative control, cells were not stimulated, only the peptide diluent (DMSO) was included. As a positive control, cells were stimulated with a polyclonal stimulant, staphylococcal enterotoxin B (SEB). Cells expressing IFN-γ and/or IL-2 and/or CD154 was the primary immunogenicity endpoint for CD4+ T cells and cells expressing IFN-γ was the primary immunogenicity endpoint for CD8+ T cells. The overall response to SARS-CoV-2 was defined as the sum of the background-subtracted responses to each of the individual pools. A sample was considered positive for CD4+ or CD8+ T cell responses to SARS-CoV-2 if any of the CD4+ or CD8+ T cell responses to the individual peptide pool stimulations was positive. Positivity was determined using MIMOSA ^39^. The total number of CD4+ T cells must have exceeded 10,000 and the total number of CD8+ T cells must have exceeded 5,000 for the assay data to be included in the analysis.

## QUANTIFICATION AND STATISTICAL ANALYSIS

### Binding and neutralizing antibody responses

Mixed effects exponential and power law models were used to analyze waning of antibody (day 42 to day 263 post symptom onset). For binding antibody analyses, antibody (Ab) was natural log transformed, yielding linear equations of the form ln(Ab)=a+b*(day-42) and ln(Ab)=a+b*ln(day/42) for the exponential and power law models, respectively, and fit using the lmer function (lme4 package) in R. Models included population level fixed effects and individual level random effects for intercept and slope and covariance between the random effects. Simplified models – with random effects only for intercept – were also fit. Neutralization antibody data were analyzed in Monolix (Lixoft). For analysis in Monolix, the exponential and power law models were formulated as ordinary differential equations, dAb/dt=k*Ab and dAb/dt=k*Ab/t, respectively, with antibody at day 42 lognormally distributed and lognormal multiplicative error. Neutralization titers < 20 were treated as left censored. For comparison of models, difference in Akaike information criterion (ΔAIC) > 4 was considered statistically Δ significant. Models (in R and Monolix) were fit using maximum likelihood. To account for repeated measures, correlations between antibody binding levels and neutralization titers were calculated using a repeated measures correlation (rmcorr package) in R ^40^.

### B cell responses

We considered linear mixed effects models for B cell response, *y_ij_* as a function of *t_ij_*, the, *j*^th^ time since symptom onset for the, *i*^th^individual, with random effects for intercept and slope and *t_ij_* > 30 days for all *i, j*

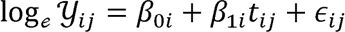

where 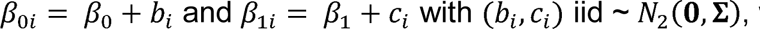, with

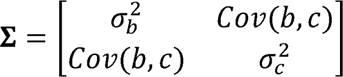

and σ_*b*_^2^ and σ_*c*_^2^ are the between-person variation in the intercept and slope of log B cell responses respectively, *Cov(b, c)* is the covariance between the intercept and slope, and ε_*i,j*_ iid ∼ *N*(0, σ^2^). The random effects, and, are each assumed to be independent for different individuals and the within-individual errors are assumed to be independent for different *i, j* and to be independent of the random effects. The function lme from the R package nlme was used to fit the models.

### T cell responses

Longitudinal analyses of CD4+ and CD8+ T cell responses were performed for individuals with a positive response for at least one time point 30 days after symptom onset. The MIMOSA (Mixture Models for Single-Cell Assays) ^39^ model incorporated cell count and cell proportion information to define a positive CD4+/CD8+ T cell response by ICS by comparing peptide pools stimulated cells and unstimulated negative controls.

This method assumed a common distribution for cytokine positive CD4+/CD8+ T cells in stimulated and unstimulated samples in non-responders, resulting in paired differences that were zero on average. In contrast, for responders, the distribution of the proportion of cytokine positive cells for stimulated samples was assumed to be greater than for unstimulated samples, resulting in paired differences that were greater than zero on average. The MIMOSA method modeled this structure through a Bayesian hierarchical mixture model framework. One component (or distribution) of the model represented the responders, and the other component modeled the non-responders. The parameters defining these distributions, as well as the probabilities that each ICS response was either a responder or non-responder, were estimated from the observed data. This sharing of information across SARS-CoV-2 responders and non-responders increased the sensitivity and specificity to make positivity calls ^41^. Responses with probability of response > 0.999 were considered positive responders.

We considered nonlinear mixed effects models for T cell response, *y*_*i,j*_, as a function of *t*_*i,j*_, the *j*^th^ time since symptom onset for the, *i*^th^ individual, with random effects for intercept and slope and > 30 days for all *i,j*

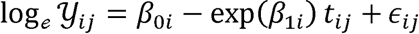

where 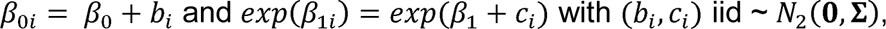, with

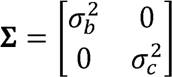

and σ_*b*_^2^ and σ_*c*_^2^ are the between-person variation in the intercept and slope of log T cell responses respectively, and ε_*i,j*_ iid ∼ *logNormal(0, σ)*. The random effects, and, are each assumed to be independent for different individuals and the within-individual errors are assumed to be independent for different *i, j* and to be independent of the random effects. The function nlme from the R package nlme was used to fit the models. Diagnostic plots of residuals were examined to assess validity of the model assumptions. Age at enrollment, gender, and disease severity (WHO score > 4) were included as covariates in the mixed effects models to assess their association with each immune response.

Individual-level estimates at days 30 (T and B cell responses), day 42 (binding and neutralizing antibody responses) and day 180 (all responses) were obtained from the mixed effects models described above. Spearman rank correlations, Wald-based two-sided 95% confidence intervals and p-values were reported.

Generalized estimating equations (GEE), with an independence working covariance matrix, were used to confirm the results of the covariate assessments for B and T cell responses from the mixed effects models. Two-tailed P-values based on the robust standard error estimates for the covariate coefficients were consistent with the corresponding two-tailed P-values for the covariate associations from the mixed effects models.

All tests were two-sided and P-values < 0.05 were considered statistically significant unless otherwise noted. Details of specific statistical analyses can be found in the Results section and in the Figure legends.

## KEY RESOURCES TABLE

| REAGENT or RESOURCE | SOURCE | IDENTIFIER |
| --- | --- | --- |
| **Antibodies** |  |  |
| Mouse Anti-Human CD3/BV510 | BD Biosciences | 564713; RRID:AB_2738909 |
| Mouse Anti-Human CD14/BV510 | BD Biosciences | 563079; RRID:AB_2737993 |
| Mouse Anti-Human CD56/BV510 | BD Biosciences | 563041; RRID:AB_2732786 |
| Mouse Anti-Human CD19/BUV395 | BD Biosciences | 563549; RRID:AB_2738272 |
| Mouse Anti-Human CD20/BUV737 | BD Biosciences | 612849; RRID:AB_2870169 |
| Mouse Anti-Human CD21/PE-Cy7 | BD Biosciences | 561374; RRID:AB_10681717 |
| Mouse Anti-Human CD27/BV605 | BD Biosciences | 302830; RRID: AB_2561450 |
| Mouse Anti-Human CD38/BB700 | BioLegend | 566445; RRID:AB_2744375 |
| Mouse Anti-Human IgA/VioBlue | Miltenyi Biotec | 130-114-005; RRID:AB_2733958 |
| Mouse Anti-Human IgD/BV650 | BD Biosciences | 740594; RRID:AB_2740295 |
| Mouse Anti-Human IgG/BV786 | BD Biosciences | 564230; RRID:AB_2738684 |
| Mouse Anti-Human IgM/PE-Dazzle 594 | BioLegend | 314530; RRID:AB_2566483 |
| Streptavidin (PE) | Invitrogen | S21388 |
| Streptavidin (AF488) | Invitrogen | S32354; RRID:AB_2315383 |
| Streptavidin (AF647) | Invitrogen | S32357 |
| Live/Dead Fixable Aqua Stain | Invitrogen | L34957 |
| Fixable Viability Dye/eFluor 450 | Invitrogen | 65-0863 |
| Mouse Anti-Human CD14/BUV661 | BD Biosciences | 741684; RRID:AB_2868407 |
| Mouse Anti-Human CD19/BUV563 | BD Biosciences | 612916; RRID:AB_2870201 |
| Mouse Anti-Human CD16/BV570 | BioLegend | 302036; RRID:AB_2632790 |
| Mouse Anti-Human CD56/BV750 | BioLegend | 362556; RRID:AB_2801001 |
| Mouse Anti-Human CD3/APC-Fire750 | BioLegend | 300470; RRID:AB_2629689 |
| Mouse Anti-Human CD4/BV480 | BD Biosciences | 566104; RRID:AB_2739506 |
| Mouse Anti-Human CD8/BUV805 | BD Biosciences | 612889; RRID:AB_2833078 |
| Mouse Anti-Human CD197(CCR7)/BV605 | BioLegend | 353224; RRID:AB_2561753 |
| Mouse Anti-Human CD45RA/BUV496 | BD Biosciences | 750258; RRID:AB_2874456 |
| Mouse Anti-Human CD25/BV650 | BD Biosciences | 563719; RRID:AB_2744337 |
| Rat Anti-Human FOXP3/PE-Cy5.5 | Invitrogen | 35-4776-42; RRID:AB_11218682 |
| Mouse Anti-Human CD32/PE-Dazzle | BioLegend | 303218; RRID:AB_2716072 |
| Mouse Anti-Human CD65/BV711 | BioLegend | 305042; RRID:AB_2800778 |
| Mouse Anti-Human CD183/PE-Cy5 | BD Biosciences | 551128; RRID:AB_394061 |
| Mouse Anti-Human CD196 (CCR6)/BV786 | BD Biosciences | 563704; RRID:AB_2738381 |
| Rat Anti-Human CD294 (CRTH2)/PE | BioLegend | 350106; RRID:AB_10900060 |
| Mouse Anti-Human IFN-g/V450 | BD Biosciences | 560371; RRID:AB_1645594 |
| Rat Anti-Human IL-2/APC | BioLegend | 500310; RRID:AB_315097 |
| Mouse Anti-Human TNF/BUV395 | BD Biosciences | 563996; RRID:AB_2738533 |
| Mouse Anti-Human IL-17A/PE-Cy7 | BioLegend | 512315; RRID:AB_2295923 |
| Rat Anti-Human IL-4/BB700 | BD Biosciences | Custom |
| Rat Anti-Human/Anti-Mouse IL-5/BB630 | BD Biosciences | Custom |
| Rat Anti-Human IL-13/BV421 | BD Biosciences | Custom |
| Mouse Anti-Human CD154 (BUV737) | BD Biosciences | 748983; RRID:AB_2873383 |
| Mouse Anti-Human Granzyme B/AF700 | BD Biosciences | 560213; RRID:AB_1645453 |
| Mouse Anti-Human Perforin/FITC | BD Biosciences | 353310; RRID:AB_2571967 |
| Mouse Anti-Human Ki-67/BB660 | BD Biosciences | Custom |
| **Bacterial and virus strains** |  |  |
| icSARS-CoV-2-mNG | Xie et al. | N/A |
| **Chemicals, peptides, and recombinant proteins** |  |  |
| SARS-CoV-2 Spike peptides | Biosynthesis | Custom |
| SARS-CoV-2 E, M, N and ORF peptides | Genscript | Custom |
| SARS-CoV-2 Spike protein (S6P) | Fred Hutchinson Cancer Research Center | Custom |
| SARS-CoV-2 RBD protein | Fred Hutchinson Cancer Research Center | Custom |
| Methylcellulose | Sigma-Aldrich | M0512-250G |
| TrueBlue Peroxidase Substrate | KPL | 5510-0050 |
| **Critical commercial assays** |  |  |
| V-PLEX COVID-19 Coronavirus Panel 2 (IgG) Kit | Meso Scale Discovery | K15369U |
| V-PLEX COVID-19 Coronavirus Panel 2 (IgA) Kit | Meso Scale Discovery | K15371U |
| V-PLEX COVID-19 Coronavirus Panel 2 (IgM) Kit | Meso Scale Discovery | K15370U |
| **Experimental models: Cell lines** |  |  |
| VeroE6 C1008 cells | ATCC | Cat# CRL-1586;<br>RRID:CVCL_0574 |
| **Software and algorithms** |  |  |
| FlowJo | BD Biosciences | V9.9.4 |
| R | R Foundation for Statistical Computing | V3.6.1 |
| GraphPad Prism | GraphPad | V7, 8 and 9 |
| Viridot | Katzelnick et al. | https://github.com/leahkatzelnick/Viridot |
| Monolix | Lixoft | MonolixSuite2019R1 |
| **Other** |  |  |
| ELISPOT reader | Immunospot | CTL ImmunoSpot S6 Universal Analyzer |

