## Supplemental Table and Figures for "Longitudinal analysis shows durable and broad immune memory after SARS-CoV-2 infection with persisting antibody responses and memory B and T cells"

### Table S1. Cohort Demographics and Baseline Characteristics (Related to STAR Methods Subject Details).

| **Characteristic** | **All (N=254)** |
| --- | --- |
| **Age, median (range)— years** | 48.5 (18-82) |
| **Female sex at birth— no. (%)** | 141 (55.6) |
| **Race or ethnic group— no. (%)** |  |
| White | 226 (89.0) |
| Hispanic or Latino | 21 (8.3) |
| Black or African American | 15 (5.9) |
| Asian | 11 (4.3) |
| Other^a^ | 7 (2.8) |
| **Median time from symptom onset to enrollment (range)— days** | 53.5 (1-203) |
| **Comorbid conditions— no. (%)** |  |
| Hypertension | 46 (18.1) |
| Obesity | 41 (16.1) |
| Chronic lung disease | 23 (9.3) |
| HIV-1 and/or autoimmune disease | 19 (7.7) |
| Type 2 diabetes mellitus | 18 (7.3) |
| Heart disease | 15 (6.0) |
| Cancer | 10 (3.9) |
| **Symptoms with initial illness— no. (%)** |  |
| Myalgia, fatigue | 231 (90.9) |
| Headache | 168 (66.1) |
| Fever | 167 (65.7) |
| Cough | 161 (63.4) |
| Loss of smell | 146 (57.5) |
| Loss of taste | 143 (56.3) |
| Shortness of breath | 108 (42.5) |
| Diarrhea | 102 (40.2) |
| Sputum production | 43 (16.9) |
| None | 9 (3.5) |
| **Disease severity (WHO Score)—no. (%)** |  |
| Mild (1-2) | 180^b^ (70.9) |
| Moderate (3-4) | 62 (24.4) |
| Severe (5-10) | 12 (4.7) |
| **Maximum number of visits—total** |  |
| 1 | 9 |
| 2 | 103 |
| 3 | 62 |
| 4 | 51 |
| 5-7 | 29 |

^a^Individuals identifying as Other included: American Indian or Alaska Native; White (n=1); Asian, Black or African American (n=1); Asian; White (n=3); Native Hawaiian or other Pacific Islander; White (n=2); ^b^6 participants had a positive Abbott SARS-CoV-2 Abbott SARS-CoV-2 IgG assay test but did not have a positive nasal SARS-CoV-2 PCR test.

**Figure S1. Modeling of antibody titer decline.** Decline of IgG antibody titers was analyzed by an exponential decay model (red) and a power law model (green) for antibodies reactive to SARS-CoV-2 antigens (A) and SARS-CoV-1 spike (B). The half-lives estimated by the exponential and power law models (C). The half-lives estimated by the power law were calculated at day 120 after symptom onset. The fold difference in IgG antibody titers to endemic coronaviruses between COVID-19 patients and pre-pandemic controls plotted over days since symptom onset (D). Related to Figure 1 and 2.

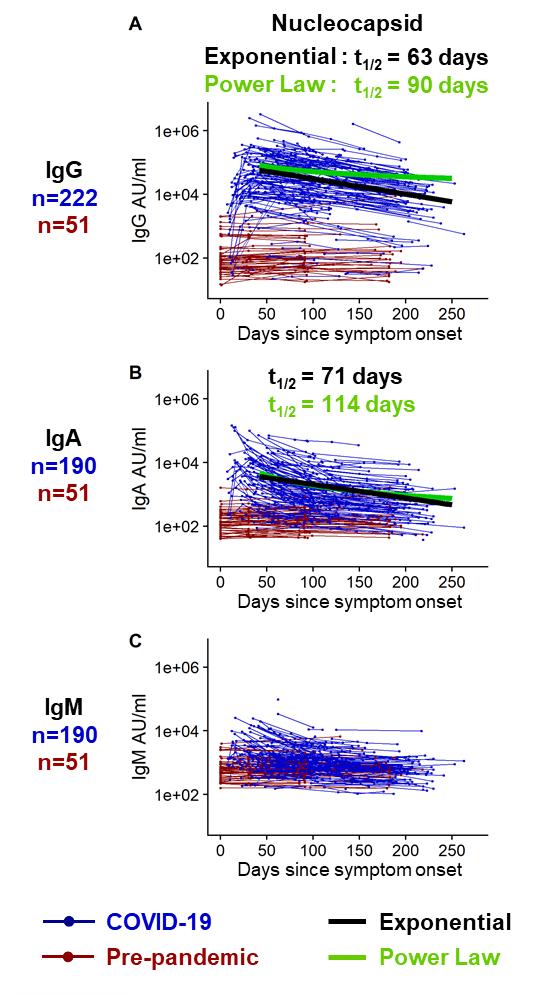

**Figure S2. Longitudinal SARS-CoV-2 nucleocapsid binding antibody responses.** IgG (A), IgA (B), and IgM (C) antibodies reactive to SARS-CoV-2 nucleocapsid were measured by an electrochemiluminescent multiplex immunoassay in triplicate and reported as arbitrary units per ml (AU/ml) as normalized by a standard curve. Longitudinal antibody titers of COVID-19 patients (in blue, n=222 COVID-19+ for IgG; n=190 COVID-19+ for IgA and for IgM) are plotted over days since symptom onset, whereas longitudinal pre-pandemic donor samples (in red, n=51 for IgG, IgA and IgM) were collected in the course of a non-SARS-CoV-2 vaccine study before 2019 and plotted over days since immunization. IgG decay curves and half-lives estimated by an exponential decay model are shown in black, whereas the decay curves and half-lives at day 120 post symptom onset estimated by a power law model are shown in green. Related to Figure 1.

| A | 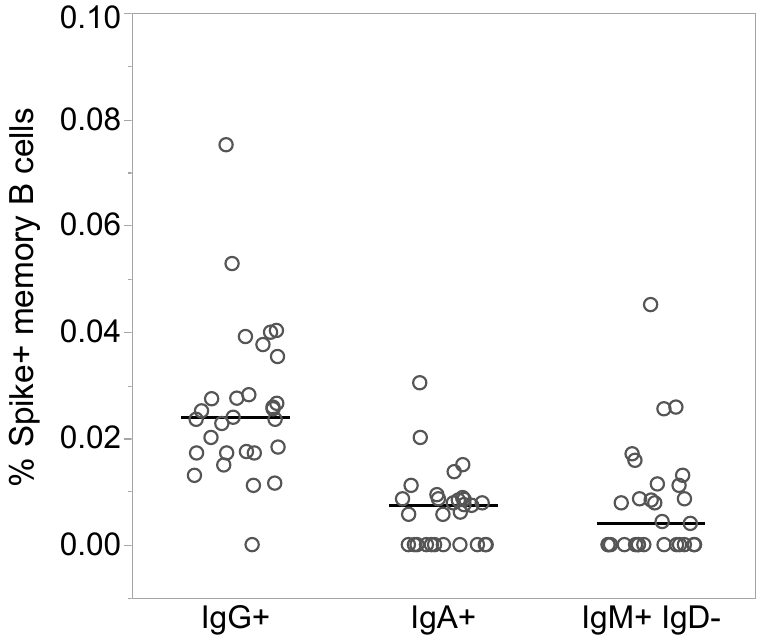 | B | 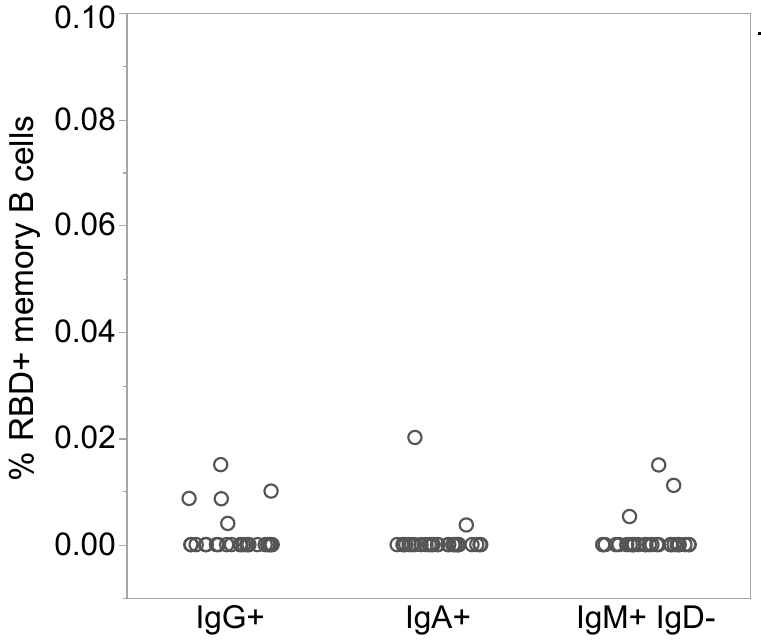 |
| --- | --- | --- | --- |
| C | 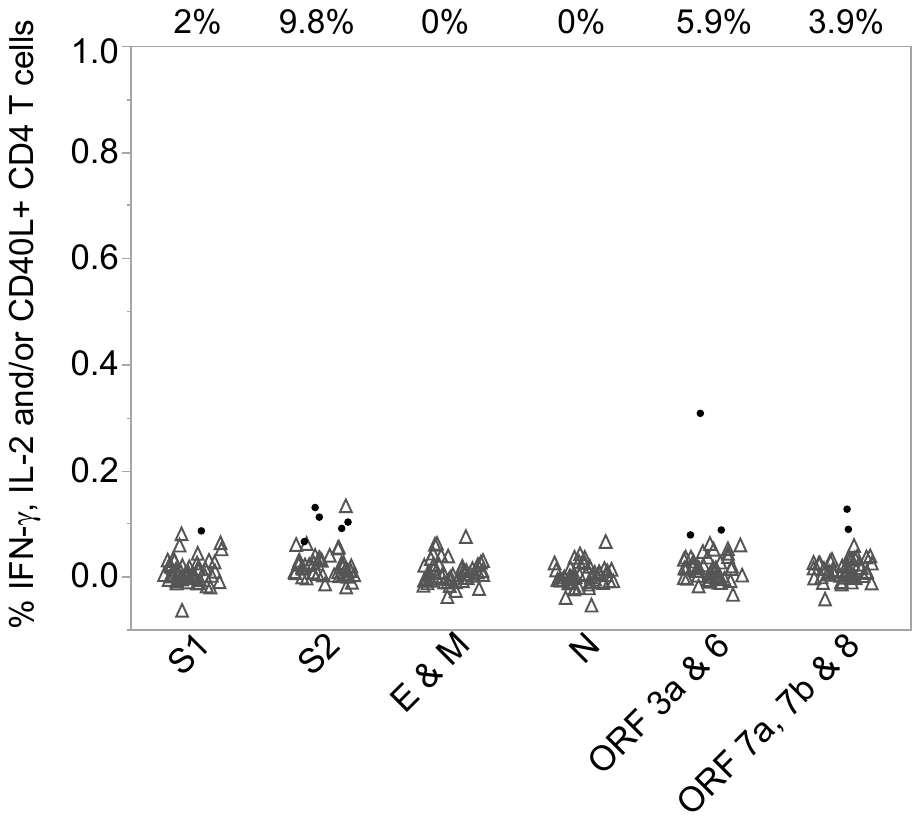 | D | 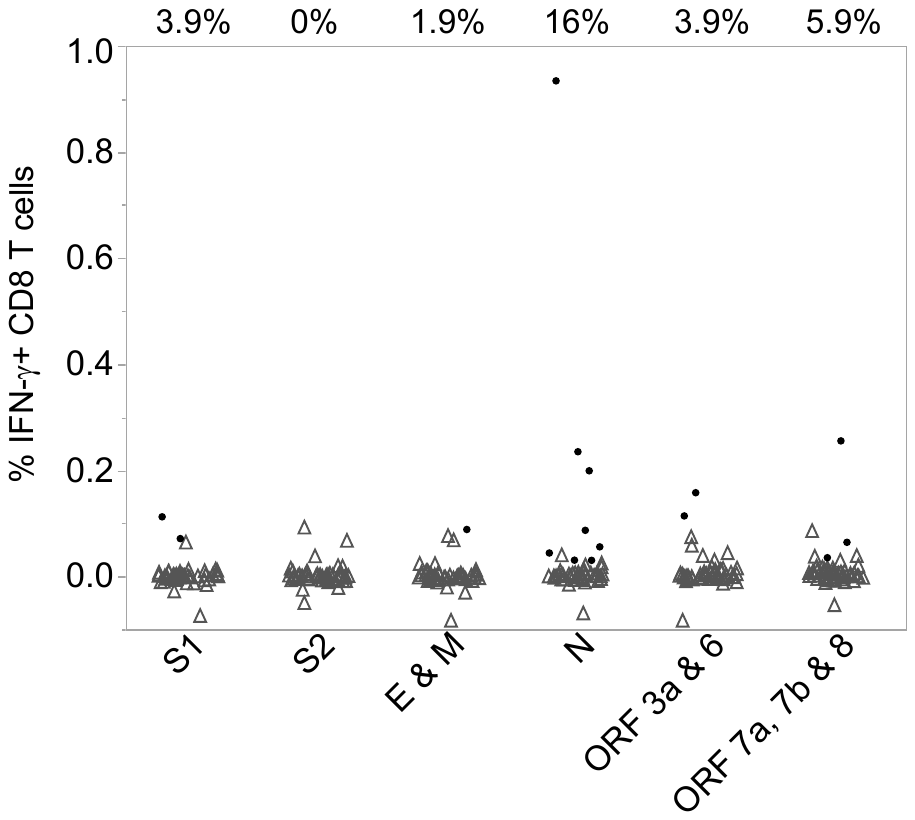 |

Figure S3. SARS-CoV-2 uninfected controls have few if any memory B and T cells recognizing SARS-CoV-2 antigens. Spike+ (A) and RBD+ (B) IgG+, IgA+ and IgM+ memory B cells in SARS-CoV-2 negative subjects are shown from PBMC collected before 2019 (n=29; tested in singlet). Line is at the median. Low frequencies of T cells recognizing SARS-COV-2 antigens are shown from donor samples not infected with SARS-CoV-2 (n=51). Background-subtracted CD4+ T cells expressing IFN-γ, IL-2 and/or CD40L (C), and IFN-γ+ CD8+ T cells (D) in response to stimulation with the SARS-CoV-2 antigens (on the x-axis) are shown. Positive T cell stimulations (as determined by MIMOSA) are indicated by a solid black circle, whereas samples that are negative are indicated by gray open triangles and the percent of positive responders are shown above the T cell graphs. Related to Figure 4, 5 and 6.

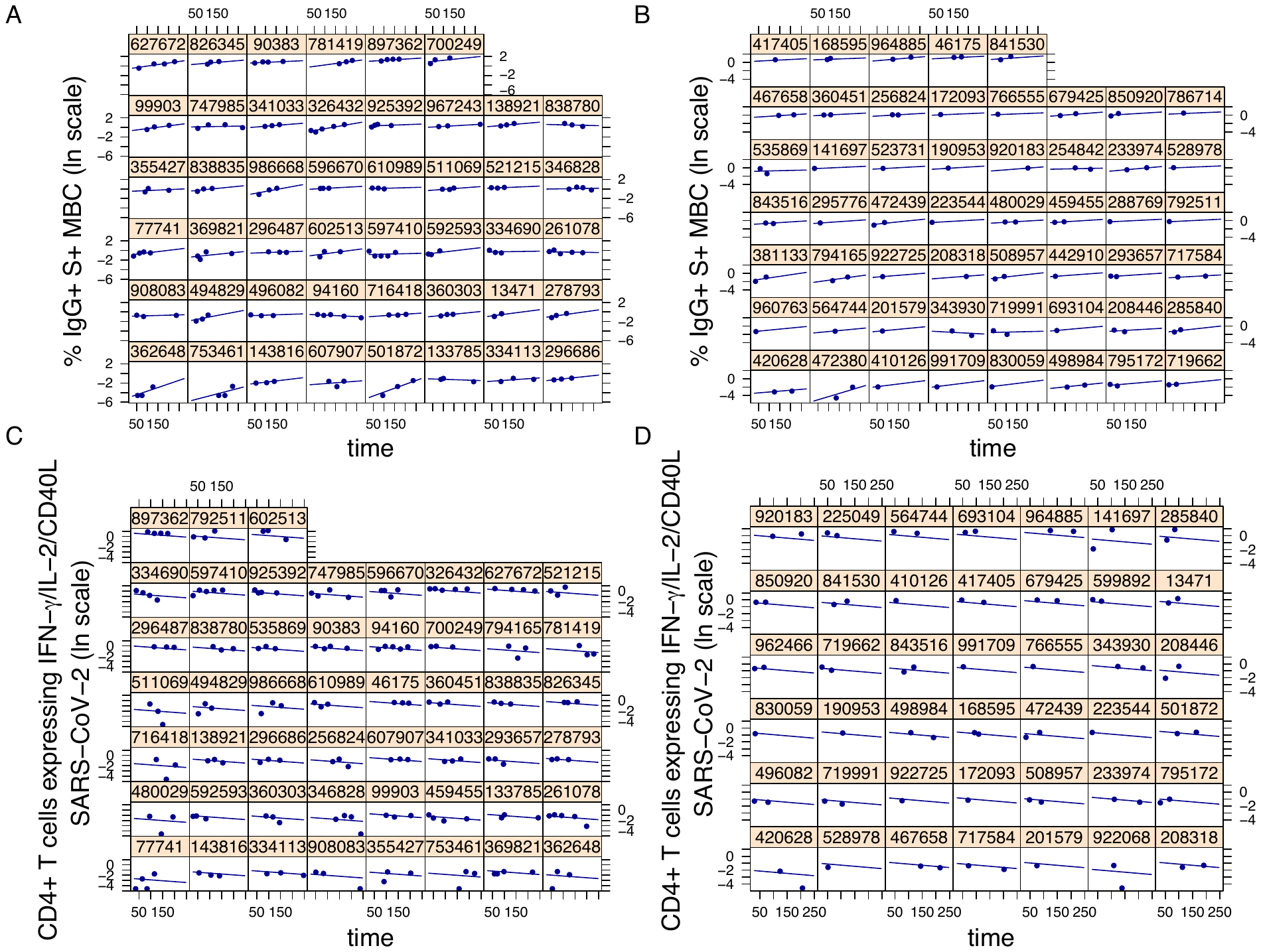

Figure S4. Representative individual-level estimates of SARS-CoV-2 B and T cell responses from 30 days post-symptom onset. Post-day 30 S+ IgG+ B cell responses (log_e_ scale) for individuals with data at 3 or more time points (A) and 1-2 time points (B) with fitted curves from a linear mixed effects model with random effects for the intercept and slope. Post-day 30 CD4+ T cell responses to SARS CoV-2 (log_e_ scale) for individuals with data at 3 or more time points (C) and 1-2 timepoints (D), with fitted curves from a nonlinear mixed effects model with random effects for the intercept and slope. The CD4+ T cell analyses only included individuals with a positive response to a least one SARS-CoV-2 antigen at one or more time points, where positive responses were determined by MIMOSA. Related to Figures 4 and 5.

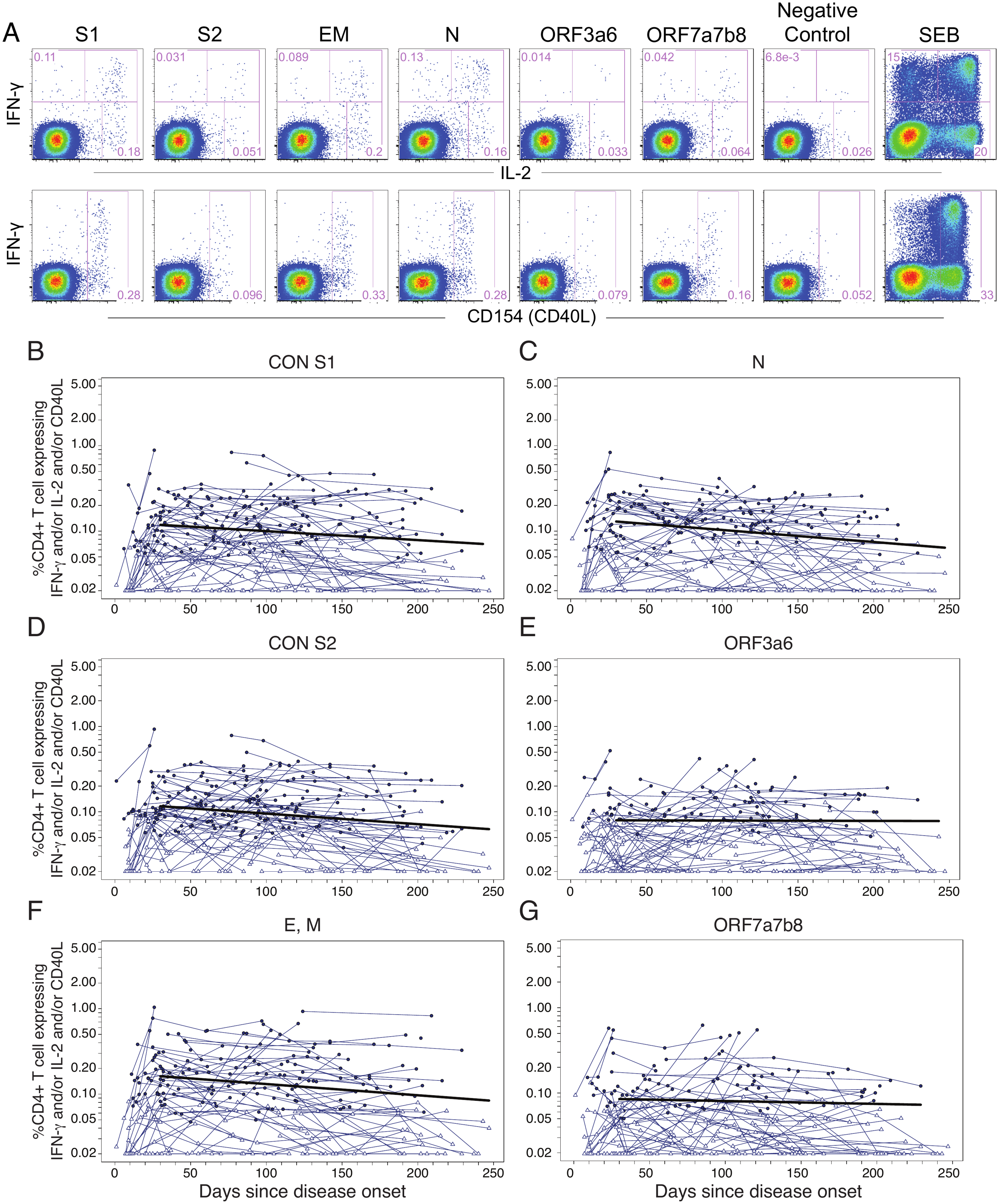

Figure S5. CD4+ T cell responses among SARS-CoV-2 convalescent subjects to individual SARS-CoV-2 peptide pools. (A) Representative SARS-CoV-2 specific CD4+ T cell responses to multiple SARS-CoV-2 antigens by intracellular cytokine staining (ICS) assay in PBMCs from a SARS-CoV-2 patient. Background-subtracted frequencies of IFN-γ+, IL-2+ and/or CD40L+ CD4+ T cells responding to: (B) S1, (C) S2, (D) envelope and membrane (EM), (E) N, (F) ORF3a and 6, (G) ORF7a, 7, and 8 (n=114; tested in single replicates). Positive responses as determined by MIMOSA are indicated by a solid circle and negative responses are indicated by open triangles. The bold black line represents the median fitted curve from a nonlinear mixed effects model of post-day 30 responses with random effects for the intercept and slope. The mixed effects models only include individuals with a positive response to the antigen(s) under consideration at one or more time points. Related to Figure 5.

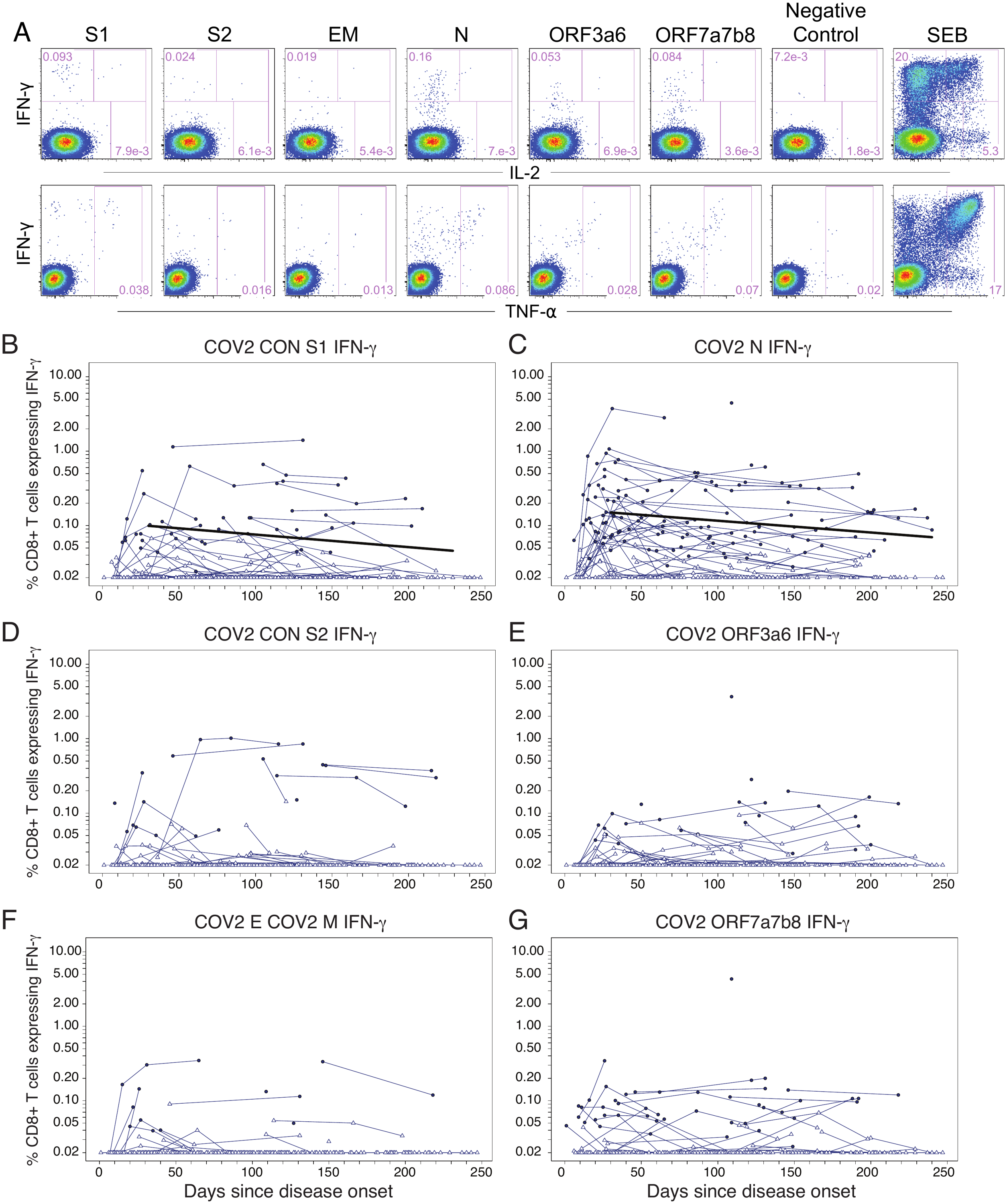

Figure S6. CD8 T+ cell responses among COVID-19 patients to individual SARS-CoV-2 peptide pools. (A) Representative SARS-CoV-2-specific CD8+ T cell responses to multiple SARS-CoV-2 antigens by intracellular cytokine staining (ICS) assay in PBMCs from a SARS-CoV-2 patient. Background-subtracted frequencies of IFN-γ+ CD8+ T cells responding to: (B) S1, (C) S2, (D) envelope and membrane (EM), (E) N, (F) ORF3a and 6, (G) ORF7a, 7, and 8 (n=114; tested in single replicates). Positive responses as determined by MIMOSA are indicated by a solid circle, and negative responses are indicated by open triangles. The bold black line represents the median fitted curve from a nonlinear mixed effects model of post-day 30 responses with random effects for the intercept and slope. The mixed effects models only included individuals with a positive response to the antigen(s) under consideration at one or more time points. Related to Figure 6.
